# Parallel Trends in an Unparalleled Pandemic Difference-in-differences for infectious disease policy evaluation

**DOI:** 10.1101/2024.04.08.24305335

**Authors:** Shuo Feng, Alyssa Bilinski

**Affiliations:** Department of Biostatistics, Brown University School of Public Health; Departments of Health Services, Policy & Practice and Biostatistics, Brown University School of Public Health

**Keywords:** Difference-in-differences, Transmission dynamics, Infectious disease models, Observational causal inference

## Abstract

Researchers frequently employ difference-in-differences (DiD) to study the impact of public health interventions on infectious disease outcomes. DiD assumes that treatment and non-experimental comparison groups would have moved in parallel in expectation, absent the intervention (“parallel trends assumption”). However, the plausibility of parallel trends assumption in the context of infectious disease transmission is not well-understood. Our work bridges this gap by formalizing epidemiological assumptions required for common DiD specifications, positing an underlying Susceptible-Infectious-Recovered (SIR) data-generating process. We demonstrate that popular specifications can encode strict epidemiological assumptions. For example, DiD modeling incident case numbers or rates as outcomes will produce biased treatment effect estimates unless untreated potential outcomes for treatment and comparison groups come from a data-generating process with the same initial infection and equal transmission rates at each time step. Applying a log transformation or modeling log growth allows for different initial infection rates under an “infinite susceptible population” assumption, but invokes conditions on transmission parameters. We then propose alternative DiD specifications based on epidemiological parameters – the effective reproduction number and the effective contact rate – that are both more robust to differences between treatment and comparison groups and can be extended to complex transmission dynamics. With minimal power difference incidence and log incidence models, we recommend a default of the more robust log specification. Our alternative specifications have lower power than incidence or log incidence models, but have higher power than log growth models. We illustrate implications of our work by re-analyzing published studies of COVID-19 mask policies.

**Significance Statement:** Difference-in-differences is a popular observational study design for policy evaluation. However, it may not perform well when modeling infectious disease outcomes. Although many COVID-19 DiD studies in the medical literature have used incident case numbers or rates as the outcome variable, we demonstrate that this and other common model specifications may encode strict epidemiological assumptions as a result of non-linear infectious disease transmission. We unpack the assumptions embedded in popular DiD specifications assuming a Susceptible-Infected-Recovered data-generating process and propose more robust alternatives, modeling the effective reproduction number and effective contact rate.

---

**T**hroughout the COVID-19 pandemic, researchers extensively studied the impact of public health interventions on disease incidence and mortality, most often with observational study designs (1). Difference-in-differences (DiD), already widely-used in health and social sciences, was one popular approach for this work. DiD assumes that treatment and non-experimental comparison groups would have moved in parallel in expectation absent the intervention (the “parallel trends assumption”). It uses this functional form assumption to impute counterfactual potential outcomes and estimate treatment effects. In the context of COVID-19, DiD was employed to evaluate policies including social distancing (2), school reopening (3), stay-at-home orders (4), and school mask mandates (5) in the United States, as well mask mandates in Germany (6) and contact tracing in England (7).

Outside of infectious disease, DiD has traditionally been used with outcomes that are expected to evolve linearly over time. By contrast, epidemiological theory predicts that infectious pathogens will spread non-linearly as a function of interactions between susceptible and infectious individuals. Mechanistic transmission dynamic models that capture these interactions are often used in epidemiology to prospectively project the impact of potential disease mitigation measures. However, these models typically rely on transporting effects from mechanistic or small-scale studies, and there are growing concerns that this may produce overly optimistic estimates of the impact of policies and programs (8). For example, randomized controlled trial estimates of the population-level reduction in transmission from mask mandates were smaller than those used to inform some transmission dynamic estimates (9). These limitations have increased interest in post-hoc policy evaluation using methods like DiD to compare outcomes in treated areas with those from similar untreated units.

However, there remains insufficient guidance on how to best account for the quasi-exponential infectious disease dynamics in DiD. Prior DiD work has noted that the parallel trends assumption may hold for some model specifications but not others (10, 11). In particular, Roth and Sant’Anna (10) demonstrated that the parallel trends assumption is sensitive to functional form unless strict conditions are met: if the population can be divided into two groups, one with random treatment assignment and another with stable distribution of the untreated potential outcomes over both pre- and post-intervention periods. Others have noted trade-offs between robustness and power and suggested applying domain-specific theory to select functional form (11).

Infectious disease transmission dynamic theory is well-developed, offering an opportunity to guide DiD specification. Nevertheless, one systematic review found that less than one-fifth of COVID-19 health policy evaluations, including many DiD applications, justified functional form or their choice of model specifications (12). To better understand the use of DiD in recent COVID-19 literature, we conducted a comprehensive review of all COVID-19 DiD analyses published in *Journal of the American Medical Association (JAMA)* network journals, *New England Journal of Medicine (NEJM), Proceedings of the National Academy of Sciences (PNAS), Nature Research* journals, *Lancet* journals, *Health Affairs*, and *Health Economics* from 2020-2022 (Appendix A). We observed considerable variation in their model specifications. Most publications (17 of the 29 papers reviewed, 59%) used incident counts or rates of cases or deaths as outcomes. Another 10 (34%) considered log-transformed incidence as the outcome measure. The remaining 2 studies (7%) specified the growth rate in log incidence as an outcome measure.

Most papers we analyzed (over 85%) did not offer specific epidemiological justification for their outcome choice. However, some researchers, particularly in economics, have cautioned against modeling incidence or mortality directly in DiD with infectious disease outcomes (3, 4). For example, Callaway and Li (4) noted that with an SIR data-generating process, the parallel trends assumption would not hold if untreated treatment and comparison trajectories differed and proposed an unconfoundedness approach to condition on pre-treatment state, in the spirit of matching on the full pre-treatment pandemic trajectory. Two papers using log growth rate as their outcome measures also motivated their specification with an SIR model (3, 13).

We extend this prior work along three key dimensions. First, we comprehensively catalog epidemiological assumptions required for DiD to recover unbiased treatment effects with different model specifications, assuming an underlying SIR process. This both summarizes the literature from an epidemiological perspective and fills in gaps by, for example, linking the log model specification to specific epidemiological parameters and expanding prior discussion of log growth models to address susceptible depletion. Second, we propose new model specifications based on epidemiological parameters. We show that these can both recover unbiased treatment effects under less strict assumptions and be applied to more complex transmission dynamic processes. Finally, we explore the power of different model specifications and highlight trade-offs involved in employing more robust specifications.

The rest of the paper proceeds as follows. Section 2 provides an overview of SIR and DiD models. In Section 3, we characterize epidemiological assumptions required for DiD to produce unbiased treatment effects assuming an underlying SIR data-generating process. We then propose alternative DiD model specifications, based on parallel trends in the effective reproduction number or effective contact rate. Section 4 explores the statistical power of different model specifications. Last, Section 5 demonstrates the implications of our findings by re-analyzing previously published work on evaluating the effect of mask policies on COVID-19 cases.

## Models

### Susceptible-Infectious-Recovered (SIR)

Transmission dynamic models assume that infectious diseases spread based on the frequency and intensity of interactions between susceptible and infectious individuals. Susceptible-Infectious-Recovered (SIR) models are a popular class of models that assume exponential growth at the start of an outbreak, with incidence declining as the susceptible population is depleted.

For this work, we posit a stochastic SIR data-generating process that incorporates randomness in transmission and, extending some prior work (e.g., Callaway and Li (4)), allows transmission intensity to vary over time, reflecting shifts in precautionary behaviors, vaccination, and variants. Given initial populations of susceptible, infectious, and recovered individuals in group *d*, {*S*_*d*,0_, *I*_*d*,0_, *R*_*d*,0_}, we assume the number of individuals in each state evolves according to the following set of equations:

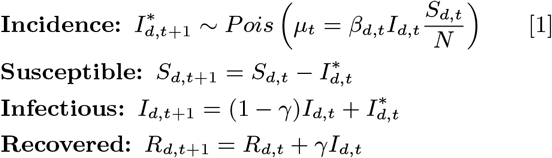

In this setup, we denote the number of incident infections for unit *d* at time *t* as 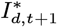 (in contrast to prevalent infections, *I*_*d*,*t*+1_). Incident infections at time *t*+1 depend on *β*_*d*,*t*_, which we denote the effective contact rate; the size of the infected population, *I*_*d*,*t*_; and the fraction of susceptible individuals, *S*_*d*,*t*_*/N* , all in the previous time step, *t*. Per standard practice, we assume incidence follows a Poisson distribution, although most results presented can accommodate other distributions with a mean of *µ*_*t*_ (4)^*^. The number of removed (recovered or dead) individuals at time *t* is *γI*_*d*,*t*_, where 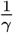 is the average length of the infectious period, equal to the generation interval in an SIR framework, i.e., the time from the infection of a primary case to a secondary infection generated.

In practice, researchers are often interested in contexts in which the susceptible population remains large relative to active and recovered infections over the period of interest, meaning that 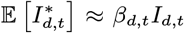. We formalize this mathematically as an “infinite susceptible population” scenario, the limit of the data-generating process as *S*_*d*,0_ → ∞ .

SIR models assume a closed and stable population of *N* individuals for every unit *d* with homogeneous mixing (i.e., *β*_*d*,*t*_*I*_*d*,*t*_*/N* to be constant across individuals). Although these assumptions are unlikely to hold exactly, they may be reasonable approximations in many applications, when researchers model outcomes at an aggregated level (e.g., counties or states). However, they may be less plausible with more granular units of analysis or when the population size is changing rapidly. We analyze this simple SIR process in closed-form; in **Extensions**, we discuss generalizations to more complex transmission dynamics.

### Difference-in-Differences (DiD)

We begin with a canonical DiD setup with two units *d* ∈ {0, 1} and two time periods *t* ∈ {*t*_1_, *t*_2_}. Assume unit 1 is treated at time *t*_2_ and unit 0 remains untreated in both time periods (14). We denote the observed outcome of interest for unit *d* at time *t* as *Y*_*d*,*t*_ and the potential outcome for unit *d* at time *t* given treatment *k* ∈{0, 1} as *Y*_*d*,*t*_(*k*). The average treatment effect on the treated (ATT) at time *t*_2_ is the expected difference in potential outcomes for unit 1 at time *t*_2_ under intervention and no-intervention scenarios:

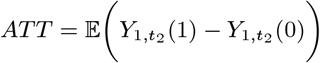

Because 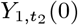 is unobservable, DiD relies on the parallel trends assumption, that the untreated potential outcomes in both units evolve in parallel from *t*_1_ to *t*_2_, i.e.,

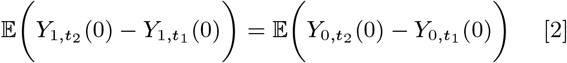

Under this assumption, we can estimate an unbiased treatment effect by solving for 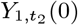 in Eq. 2 and applying sample analogs:

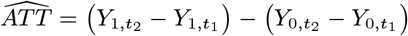

We can extend the parallel trends assumption to monotonic, continuously differentiable transformations (*g*(·)) (15):

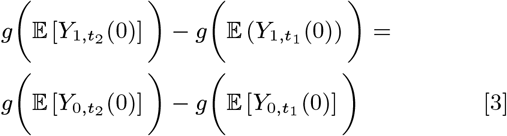

In the following sections, we establish conditions under which the transformed non-linear version of parallel trends assumption (Eq. 3) holds under different specifications of the DiD model, i.e., different *Y*_*d*,*t*_ and *g*(·). In **Extensions**, we generalize findings to DiD applications with more than two units or time periods, noting that the conditions remain largely unchanged.

### Model specifications

Assuming an underlying SIR data-generating process, different model specifications encode different epidemiological conditions required for the parallel trends assumption to hold, allowing unbiased estimation of the ATT with DiD. In this section, we derive and provide intuition for assumptions invoked across various specifications.

We summarize model specifications considered in Table 1, increasing in robustness. These include the three specifications identified in our literature review, as well as two new proposed specifications modeling the effective reproduction number and effective contact rate. Each specification implies a different interpretation of the ATT as noted in the fourth column of Table 1. As a result, we detail methods for imputing the average marginal effect on the incidence scale in **Average marginal effects**.

**Table 1.**
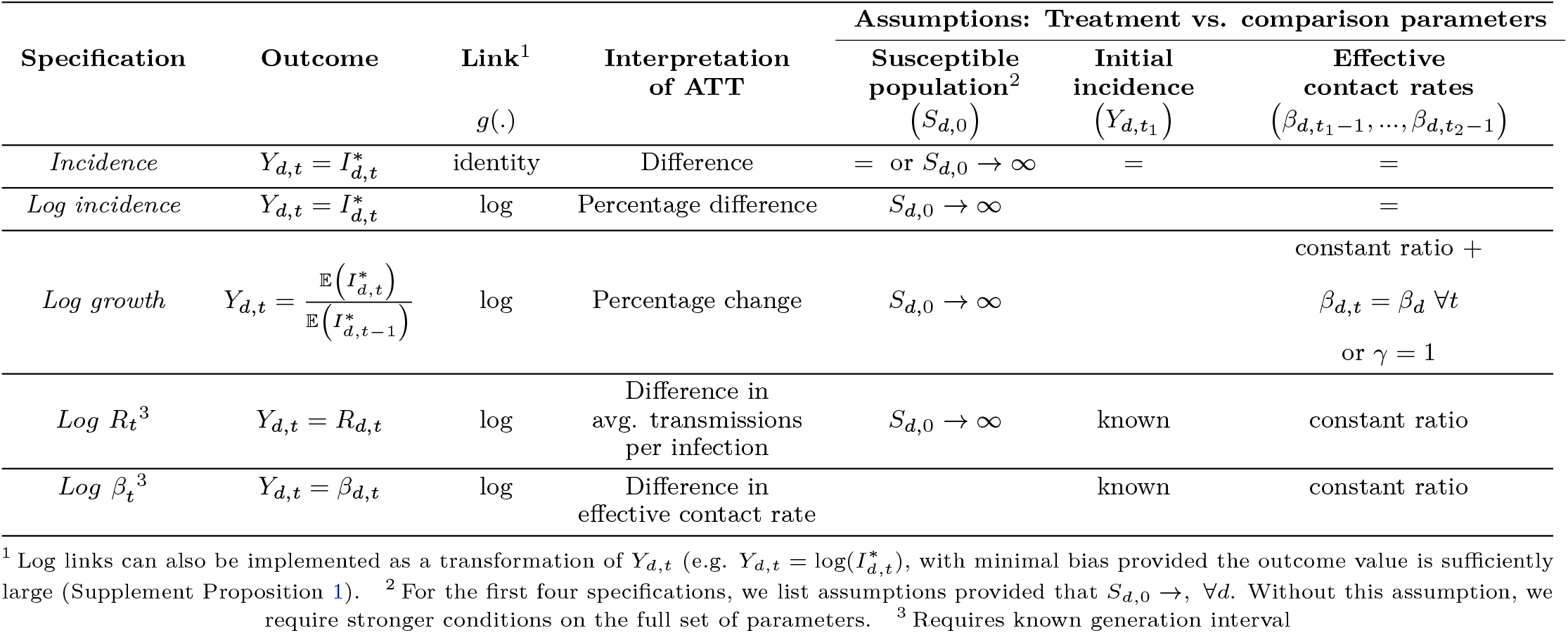
Transmission dynamic assumptions required for the parallel trends assumption, assuming an SIR data-generating process.

| Specification | Outcome | Link<sup>1</sup><br>$g(\cdot)$ | Interpretation<br>of ATT | Assumptions: Treatment vs. comparison parameters | | |
| --- | --- | --- | --- | --- | --- | --- |
| | | | | Susceptible<br>population<sup>2</sup><br>($S_{d,0}$) | Initial<br>incidence<br>($Y_{d,t_1}$) | Effective<br>contact rates<br>($\beta_{d,t_1-1}, \dots, \beta_{d,t_2-1}$) |
| *Incidence* | $Y_{d,t} = I_{d,t}^*$ | identity | Difference | $=$ or $S_{d,0} \rightarrow \infty$ | $=$ | $=$ |
| *Log incidence* | $Y_{d,t} = I_{d,t}^*$ | log | Percentage difference | $S_{d,0} \rightarrow \infty$ | | $=$ |
| *Log growth* | $Y_{d,t} = \frac{\mathbb{E}(I_{d,t}^*)}{\mathbb{E}(I_{d,t-1}^*)}$ | log | Percentage change | $S_{d,0} \rightarrow \infty$ | | constant ratio +<br>$\beta_{d,t} = \beta_d \forall t$<br>or $\gamma = 1$ |
| *Log $R_t$<sup>3</sup>* | $Y_{d,t} = R_{d,t}$ | log | Difference in<br>avg. transmissions<br>per infection | $S_{d,0} \rightarrow \infty$ | known | constant ratio |
| *Log $\beta_t$<sup>3</sup>* | $Y_{d,t} = \beta_{d,t}$ | log | Difference in<br>effective contact rate | | known | constant ratio |

<sup>1</sup> Log links can also be implemented as a transformation of $Y_{d,t}$ (e.g. $Y_{d,t} = \log(I_{d,t}^*)$ ), with minimal bias provided the outcome value is sufficiently large (Supplement Proposition 1). <sup>2</sup> For the first four specifications, we list assumptions provided that $S_{d,0} \rightarrow \infty, \forall d$ . Without this assumption, we require stronger conditions on the full set of parameters. <sup>3</sup> Requires known generation interval

For log specifications in Table 1, we assume that the parallel trends assumption is defined in terms of log (𝔼 [*Y*_*d*,*t*_]). However, 8 (or 28%) papers modeled log specifications by transforming the outcome variable, assuming parallel trends in 𝔼 [log (*Y*_*d*,*t*_)], rather than log (𝔼 [*Y*_*d*,*t*_]). Although 𝔼 [log (*Y*_*d*,*t*_)] ≠ log (𝔼 [*Y*_*d*,*t*_]) (Jensen’s inequality), the two are nearly equivalent when *Y*_*d*,*t*_ is sufficiently large (Supplement Proposition 1 and Corollary 1), and in practice, results are unlikely to be affected by this approximation.

Last, note that each model specification in Table 1 can be formulated based either on the count or rate per population of infections. In the literature reviewed (Appendix A), about half (48%) of researchers modeled rates (i.e., infections or cases per unit population) rather than numbers of infections or cases. This assumes frequency-dependent transmission, i.e., the average number of secondary infections per infectious individual remains relatively constant across different population sizes (16). For parsimony in this section, we assume that units have equal population sizes, obviating the need to scale in derivations, and results that follow could apply either to frequency-dependent or density-dependent transmission. However, if population sizes differ and frequency-dependent transmission is assumed, derivations can be adapted accordingly by scaling model specifications by population size (e.g., incidence per 100,000 population rather than incident infection numbers).

### Established model specifications

#### Incidence

As previously noted, incidence was the most popular model specification: 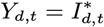 (with the identity link function). To understand epidemiological assumptions embedded in DiD with incidence outcomes, we first obtain an expression for expected incidence.

##### Proposition 1

(Expected incidence). *Assuming an SIR data-generating process (Eq. 1) with initial conditions {S*_*d*,0_, *I*_*d*,0_, *R*_*d*,0_ }, *expected incidence at time t* + 1 *can be written as:*

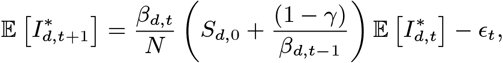

*where* 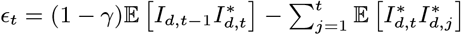 .

We derive Proposition 1 in Appendix B. Even with *t*_1_ and as adjacent time-steps, we cannot write 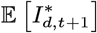 as a linear function of 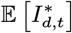. Indeed, with this data-generating process, there are not straightforward conditions under which the parallel trends assumption holds for incidence, log incidence, or log growth models other than equality in all data-generating parameters.

We therefore explore assumptions required under “infinite susceptible population” conditions (i.e., *S*_*d*,0_→ ∞ ), implying that the susceptible population is very large relative to active and recovered infections. In this case, expected incidence can be written as an iterative product of prior effective contact rates and initial infection.

##### Proposition 2

(Expected incidence (infinite susceptible population)). *Assuming an SIR data-generating process (Eq. 1), with initial conditions* {*S*_0_, *I*_0_, *R*_0_}, *for t* ≥ 1,

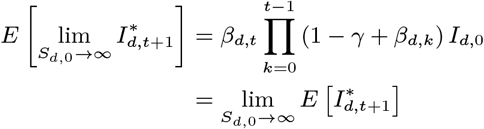

We then solve for conditions under which the “infinite susceptible population” parallel trends assumption holds as *S*_1,0_, *S*_0,0_ → ∞. Because 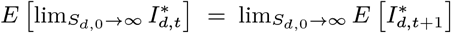 , and *g*(·) is continuous, we write 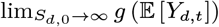 to compress notation; because units are assumed independent, taking the limit as *S*_1,0_, *S*_0,0_→ ∞ is equivalent to taking the corresponding limit on each unit.

##### Proposition 3

(Parallel trends: Incidence (infinite susceptible population)). *Assuming an SIR data-generating process (Eq. 1) and an incidence model specification* 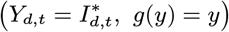, *the “infinite susceptible population” parallel trends assumption (Eq. 3) holds between t*_1_ *and t*_2_ *under the following conditions:*

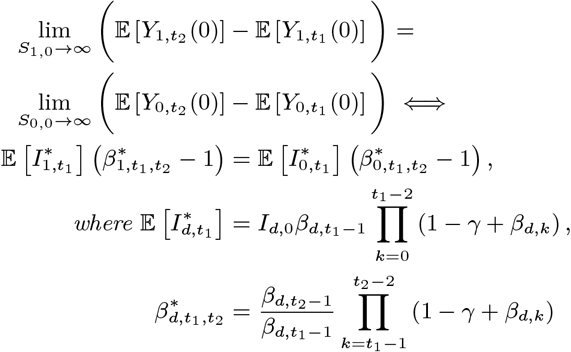

Proposition 3 follows from substituting results of Proposition 2 into Eq. 3 (proof in Appendix B). This result highlights that the parallel trends assumption imposes strong conditions on underlying transmission dynamics, even assuming an “infinite susceptible population. Proposition 3 holds when groups start with the same expected incidence 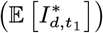 at time *t*_1_ and continue to grow with a set of time-varying effective contact rates (*β*_*t*_) and generation interval (*γ*) that produce equal 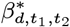. (Parallel trends over time would require equality across groups in this set of effective contact rates). We illustrate these assumptions the first row of Figure 1, where trends are only parallel (noted by a checkmark) when all input parameters are equal, and as a result expected incidence trajectories match, between treatment and comparison groups.

**Fig. 1.**
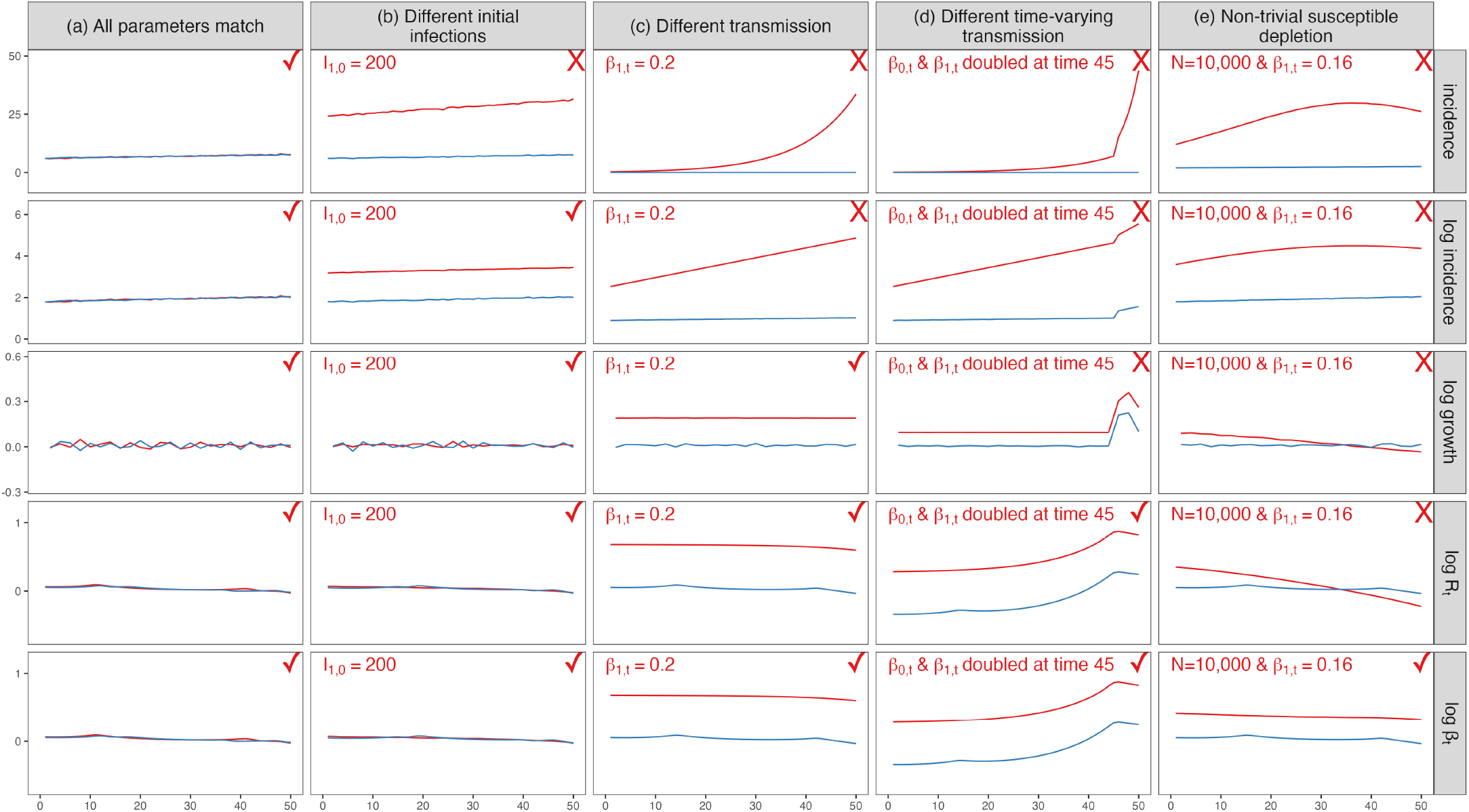
Comparison of different model specifications. We vary the choice of outcome variable across rows and the underlying data-generating process across columns. We simulate data from an SIR model and average outcomes over 1000 draws. Lines indicate the average outcome value for two units. Assume unit 0 is graphed in blue, and unit 1 is graphed in red. There are no treatment effects in any scenario; therefore plots display untreated potential outcomes. If trends are parallel (checkmark), then DiD would estimate unbiased treatment effects with the corresponding model specification. Baseline parameters are *N* = 10^8^, *I*_0_ = 50, 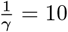, *β*_*t*_ = 0.105 ∀ *t*. (a) “All parameters match” matches all parameters between units for all *t*. (b) “Different initial infections” changes the initial number of infections for the unit graphed in red to *I*_1,0_ = 100. (c) “Different transmission” sets *β*_1,*t*_ = 0.2. (d) “Different time-varying transmission” sets *β*_1,*t*_ = 0.2 for the treated unit and doubles both *β*_0,*t*_ and *β*_1,*t*_ for time *t* ≥ 45. (e) “Non-trivial susceptible depletion” sets a smaller population size *N* = 10^4^ for both units and sets *β*_1,*t*_ = 0.160.

Proposition 3 also implies that traditional DiD diagnostics like event study plots may be misleading for assessing the plausibility of parallel trends assumption. In contrast to traditional DiD which allows for level differences, this specification requires evidence of equal epidemic trajectories. Furthermore, visual inspections may be misleading at the start of a new wave of disease, as groups may initially appear similar, but increasingly diverge as a wave accelerates even absent a treatment effect (Figure 1, panel 1(b)).

#### Log incidence

The second most common model specification was log incidence, 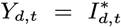 with *g*(·) = log(·), often motivated by the idea that disease transmission can be exponential or nearly exponential (17, 18).

##### Proposition 4

(Parallel trends: Log incidence (infinite susceptible population)). *Assuming an SIR data-generating process (Eq. 1) and a log incidence model specification* 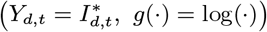, *the “infinite susceptible population” parallel trends assumption (Eq. 3) holds between t*_1_ *and t*_2_ *under the following conditions:*

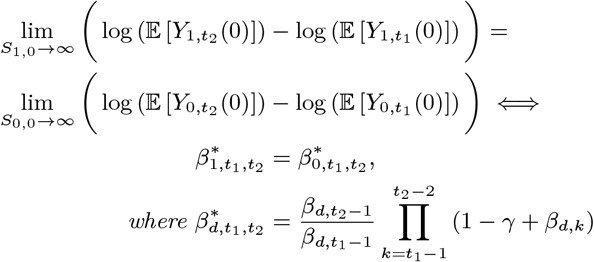

The derivation for Proposition 4 follows similar logic as that of Proposition 3 (Appendix B). Per Proposition 4, with an “infinite susceptible population,” when log incidence is used as outcome, the parallel trends assumption no longer imposes restrictions on expected incidence at time *t*_1_. Nevertheless, it still requires an equal product of effective contact rates in both units between *t*_1_ and *t*_2_. We illustrate these conditions in the second row of Figure 1.

#### Log growth

The third approach identified in our literature review (“log growth”) modeled the change in log incidence over adjacent time steps. Because this ratio is undefined in the context where 0 is in the support of 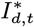, we model this as 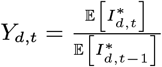, with *g*(·) = log(·).^†^

##### Proposition 5

(Parallel trends: Log growth (infinite susceptible population)). *Assuming an SIR data-generating process (Eq. 1) and a log growth model specification* 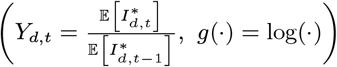 , *the “infinite susceptible population” parallel trends assumption (Eq. 3) holds between t*_1_ *and t*_2_ *under the following conditions:*

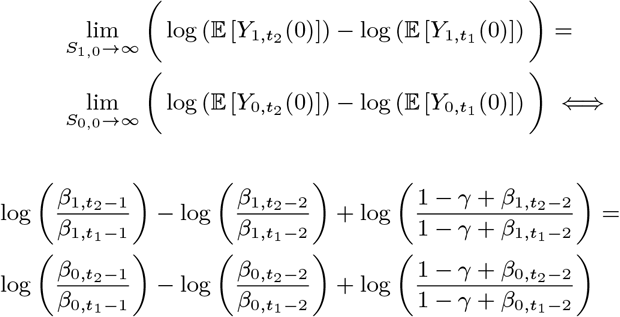

The implications of this condition are less straightforward than those of prior specifications. First, the log growth specification again imposes no requirements on expected incidence at *t*_1_. Furthermore, although these results invoke the “infinite susceptible population” assumption, this is less restrictive in the log growth specification, a result of dividing incidence at adjacent time-steps.

In the special case where the length of generation interval is equal to the time step (*γ* = 1, i.e., no intergenerational compounding)^‡^, Proposition 5 becomes:

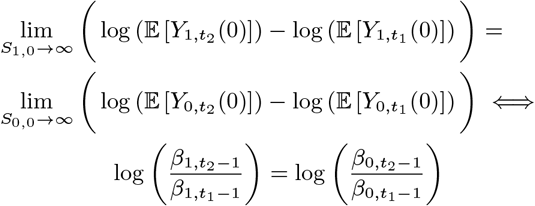

Effective contact rates could then be allowed to differ between groups and vary over time, providing that their ratios remain constant across groups. In practice, this means that unbiased treatment effect estimation could be achieved when units differ in terms of baseline mask mandates or vaccination coverage, provided this ratio remains unchanged over the study period. However, this condition imposes restrictions on both the generation interval (*γ*) and susceptible depletion (Figure 1, panels 3(c) through 3(e)).

### Proposed model specifications

Of the specifications characterized above, log growth is the most flexible, allowing the effective contact rate to differ across groups by a constant ratio under certain conditions. However, these may be too narrow for many applications. We therefore propose two alternatives that allow us to relax assumptions further by drawing on common epidemiological quantities: the log of the effective reproduction number (*R*_*t*_) and of the effective contact rate (*β*_*t*_).

#### Log effective reproduction rate (R_τ_)

The effective reproduction number, traditionally denoted *R*_*t*_,^§^ measures the average number of secondary infections caused by each individual infection over a time interval traditionally denoted *t*. This quantity is commonly used to understand the risk of exponential spread in a population (19), accounting for contact patterns, precautionary behaviors, and immunity (20). It is often benchmarked against a value of 1: if *R*_*t*_ *<* 1, incident infections will decrease over time, and the goal of policies is often to maintain *R*_*t*_ below 1 (18, 19, 21).

In this section, we consider modeling log(*R*_*t*_) as the outcome variable in DiD and demonstrate that this model specification further relaxes the assumptions about the underlying transmission dynamics. Although there are a few possible formulations of the effective reproduction number based on *β*_*t*_ (22), we use the popular cohort-based definition (21), which can be more easily mapped to intervention timing than “instantaneous” approaches (e.g., Cori et al. (19)).

##### Proposition 6

(Cohort definition of *R*_*t*_). *Assume that the effective reproduction number is measured over a generation interval of length* 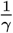 *for the cohort* 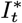 *becoming infectious at time t. We define the cohort effective reproduction number:*

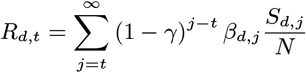

With this link (proof in Appendix B), we can characterize the conditions required for the parallel trends assumption to hold when the outcome specification is log *R*_*d*,*t*_.

##### Proposition 7

(Parallel trends: Log *R*_*t*_ (infinite susceptible population)). *Assuming an SIR data-generating process (Eq. 1), log-transformed effective reproduction number model specification* (*Y*_*d*,*t*_ = log (*R*_*d*,*t*_) , *g*(·) = log(·)), *the “infinite susceptible population” parallel trends assumption (Eq. 3) holds if and only if*

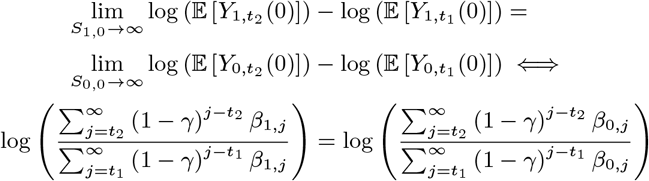

Per Proposition 7, the log *R*_*t*_ specification requires only a constant ratio between effective contact rates in treatment and comparison groups during time-steps included in the *R*_*t*_ estimate. With a short generation interval, aggregation presents few practical concerns; with longer generational intervals, researchers may be wary about spillover between treatment and comparison timings.

Like other specifications discussed, this specification also is sensitive to differential susceptible depletion. For small populations or infectious diseases with intense transmission, the susceptible population may deplete quickly and may do so differentially across treated and comparison groups if they start with different susceptible fractions. In this case, DiD using log *R*_*t*_ as an outcome would produce biased treatment effect estimates (Figure 1, panel 4(e)). In the context of COVID-19, population susceptibility often declined rapidly during major waves, underscoring the value of having an model specification robust to non-trivial susceptible depletion.

#### Log effective contact rate (β_τ_)

Although the effective reproduction number is more often modeled in applied epidemiology, the effective contact rate may be a more robust outcome. Proposition 8 formalizes the conditions required for the parallel trends assumption to hold when using a log *β*_*t*_ model specification.

##### Proposition 8

(Parallel trends: Log *β*_*t*_). *Assuming an SIR data-generating process (Eq. 1) and a log-transformed effective reproduction number specification* (*Y*_*d*,*t*_ = log (*β*_*d*,*t*_, *g*(·) = log(·))) *the parallel trends assumption (Eq. 3) holds if and only if*

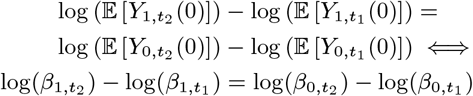

Proposition 8 follows from substituting *Y*_*d*,*t*_ = *β*_*d*,*t*_ into the log-transformed parallel trends assumption as defined in Eq. 3. This specification simplifies the assumptions imposed by *R*_*t*_, requiring only a constant ratio on *β*_*t*_ and is not sensitive to susceptible depletion.

### Estimation

#### R_τ_ and β_τ_

Our two proposed outcomes, *R*_*t*_ and *β*_*t*_, are not straightforward transformations of incident infections, but can be obtained via maximum likelihood estimation assuming an SIR model. For example, under the SIR data-generating process (Eq. 1), we can obtain a maximum likelihood estimate of *β*_*t*_:

##### Proposition 9

(Estimation of *β*_*d*,*t*_). *Assuming an SIR data-generating process (Eq. 1), with* 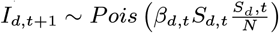, *the maximum likelihood estimator of β*_*d*,*t*_ *is:*

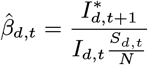

Beyond SIR models, we can also estimate *R*_*t*_ and *β*_*t*_ assuming more complex data-generating processes, another benefit over incidence, log incidence, or log growth specifications discussed further in **Extensions**.

#### ATTs

We can estimate ATTs of interest through regression:

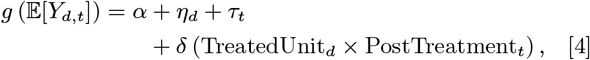

where *Y*_*d*,*t*_ refers to the chosen model specification, *α* denotes the intercept, *η*_*d*_ denotes the unit fixed-effect for each unit *d*, and *τ*_*t*_ denotes the time fixed effect for each time period TreatUnit_*d*_ and PostTreatment_*t*_ are binary variables indicating whether unit *d* is ever treated and whether the time period *t* is post-intervention, respectively. With this setup, *δ* is the ATT of interest. When *g*(·) is the identity, we use OLS, and for a log link, we use Poisson regression. (As mentioned above, we could often nearly equivalently model 𝔼 (log[*Y*_*d*,*t*_]) with OLS, but Poisson regression circumvents the issues of managing zeros in the outcome variable with a log transformation (10)). We use the wild score bootstrap for inference (Appendix G) (23, 24).

### Average marginal effects

When using non-incidence model specifications, estimated regression coefficients may not provide useful interpretations beyond their signs. To improve interpretability, we convert coefficients from non-incidence specifications back to the incidence scale by calculating their average marginal effects (AME), enabling comparison across different model specifications.

For log incidence and log growth models, we first estimate untreated potential outcomes for the treated group using the observed outcome trajectories in the treated units and the estimated ATT and convert each to the incidence scale. We then compute the difference between potential and observed outcomes in the post-intervention period, and average the differences over all units to obtain the AME. For log *R*_*t*_ and log *β*_*t*_ models, we estimate the potential outcomes for each unit by simulating infections from an infectious disease transmission model. We simulate the treated and untreated potential trajectories for treated units 1000 times and calculate the difference in the post-intervention period to estimate AMEs. Detailed algorithms are provided in Appendix E.

### Extensions

#### Generalizations of SIR models

In practice, epidemiologists rarely assume simple SIR data-generating processes, in favor of models that capture more granular transmission dynamics. For example, many models introduce an additional exposed state (Susceptible-Exposed-Infectious-Recovered [SEIR]), which tracks exposed but not yet infected individuals (Appendix F) (25). Models can also be adapted to include heterogeneity across subgroups or to agent-based frameworks that allow heterogeneity in individual agents.

Our proposed model specifications involving the effective reproduction number and the effective contact rates remain conceptually meaningful quantities and can still be estimated assuming more complex transmission dynamics. Wallinga and Teunis’s cohort-based *R*_*t*_ estimator can accommodate a wide range of underlying data-generating processes, producing an unbiased estimate when a directed network for infectious disease transmission is defined and the generation interval is known or can be estimated (21). When the time-step is approximately equal to the generation interval, estimates of *R*_*t*_ can be converted to *β*_*t*_ by dividing by the susceptible population fraction. (For COVID-19 applications detailed below, we assume a generation time of approximately a week and model *R*_*t*_ at weekly intervals). More complex approximation of a time-varying *β*_*t*_, rather than assuming constant *β*_*t*_ over a generation interval, may be needed when generation intervals are longer.

#### Time-step aggregation

When a disease is relatively rare, researchers may choose to aggregate data over time above the level of the infectious disease process timestep (e.g., aggregating days into weeks or months). In this case, we require the aggregated parallel trends assumption:

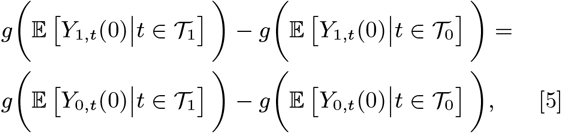

where *𝒯*_0_ and *𝒯*_1_ are the collections of pre- and post-treatment time periods. Here for Eq. 5 with an identity or a log link to be true, it is sufficient to assume that the original parallel trends assumption (Eq. 3) holds for every two time periods *t*_1_ and *t*_2_ such that 1 ≤ *t*_1_ ≤ *T*_0_ *< t*_2_ ≤ *T* , where *T*_0_ is the time of intervention. This result follows from the linearity of expectation for the identity link; for a log link, we derive the condition in Supplement Proposition 2.

#### Multiple units and time periods

In practice, researchers often have more than 2 periods and more than 2 units in their study population. We can extend the parallel trends assumption (Eq. 3) to both multiple units and multiple time periods:

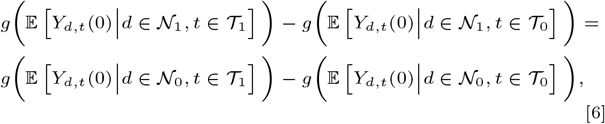

where *𝒩*_1_ and *𝒩*_0_ denote the collections of treated and control units and *𝒯*_0_ and *𝒯*_1_ are the collections of pre- and post-treatment time periods. In this context, we can estimation procedures following those outlined in **Estimation**.

### Multiple pre- and post-time periods

With multiple pre- and post-time periods, we can meet the conditions for Eq. 6 with the same parallel expected trajectories condition required for time-step aggregation (Corollary 2). This translates to similar conditions required for each model specification in Propositions 3-5 and 7-8, except that conditions must hold for all time period pairs, rather than just *t*_1_ and *t*_2_.

In the special case of staggered intervention roll-out, simply averaging over the post-intervention treatment effects does not yield a sensible estimand unless additional homogeneity assumptions are imposed (14). Recently developed estimators allow for more flexible treatment effect estimation, such as those proposed by Callaway and Sant’Anna (26), Sun and Abraham (27) and Borusyak et al. (28). In general, these methods proposed ways to construct a group-time treatment effect for each treatment-adoption cohort, aggregating these group-time effects to obtain an overall ATT, and the above assumption would again suffice for unbiased effect estimation.

### Multiple treated and comparison units

With multiple treated units, Eq. 6 requires that transformed average differences between pre- and post-treatment outcomes be equal across treated and untreated units. This too follows from assuming parallel trends between pairs of treated and comparison units. Outside of this scenario (or scenarios with equal data-generating parameters for all units within treatment or comparison groups), researchers may need to exercise care in identifying appropriate groups, particularly for incidence specifications, as equal averages across data-generating parameters for treatment and comparison groups is not sufficient for equal expected incidence (Proposition 2).

## Power Analysis

Power to detect treatment effects may vary across model specifications (29). We therefore performed a simulation study to explore the performance of the five model specifications in the previous section.

### Simulation setup

With an SIR model (Eq. 1), we simulated infection trajectories in 50 units over a total time interval of 17 weeks, with an average generation interval of 10 days 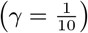. Twenty-five units (50% of all units) were randomly designated as treated units. We introduced 100 seed infections to the population at time 0 and set a constant baseline effective contact rate *β*_*t*_ ∈ { 0.100, 0.115 }. We then varied the baseline effective contact rate in treated units to be either equal to that of untreated units or 10% greater. For treated units, we introduced an intervention that changed the effective contact rate at week 9. We varied the magnitude of the treatment effect as a multiplicative factor (*δ*) on the baseline transmission rate, by setting 0.70 ≤ *δ* ≤ 1.3. The scenario *δ* = 1 corresponded to the null-effect case, i.e., no change in transmission is introduced in the treatment group. We also varied the population size in each unit, *N* ∈ { 5000, 10000 }. Simulation specifications are also summarized in Table A1.

We estimated the five outcomes outlined in Table 1. For *R*_*t*_, we used the Wallinga-Teunis estimator and divided estimates by the susceptible fraction to estimate *β*_*t*_. After estimating these outcome, we excluded the first 5 weeks of data for all specifications because previous studies have noted *R*_*t*_ estimates are often biased at the start of a time series (22).^¶^ We also discarded the last 5 weeks of data to account for a similar bias due to delayed reporting (21).

We estimated treatment effects per **Estimation** and reported the probability of rejecting the null in each scenario using confidence intervals generated by wild score bootstrap (24). These corresponded to power in the case where *δ* ≠ 1 and type I error where *δ* = 1. We used a significance level of 0.05, and all results shown are an average of 1000 simulations.

### Simulation results

Figure 2 summarizes the power (*δ* ≠ 1) and type I error (*δ* = 1) when initial infections, initial susceptible fractions, and transmission parameters are all equal between the treatment and control groups. The left and right panels differ in the scale of effect size: the left plots the effect size (*δ*) which is a multiplicative factor on the baseline effective contact rate, and the right plots the percentage change in infection due to the change in effective contact rate.

**Fig. 2.**
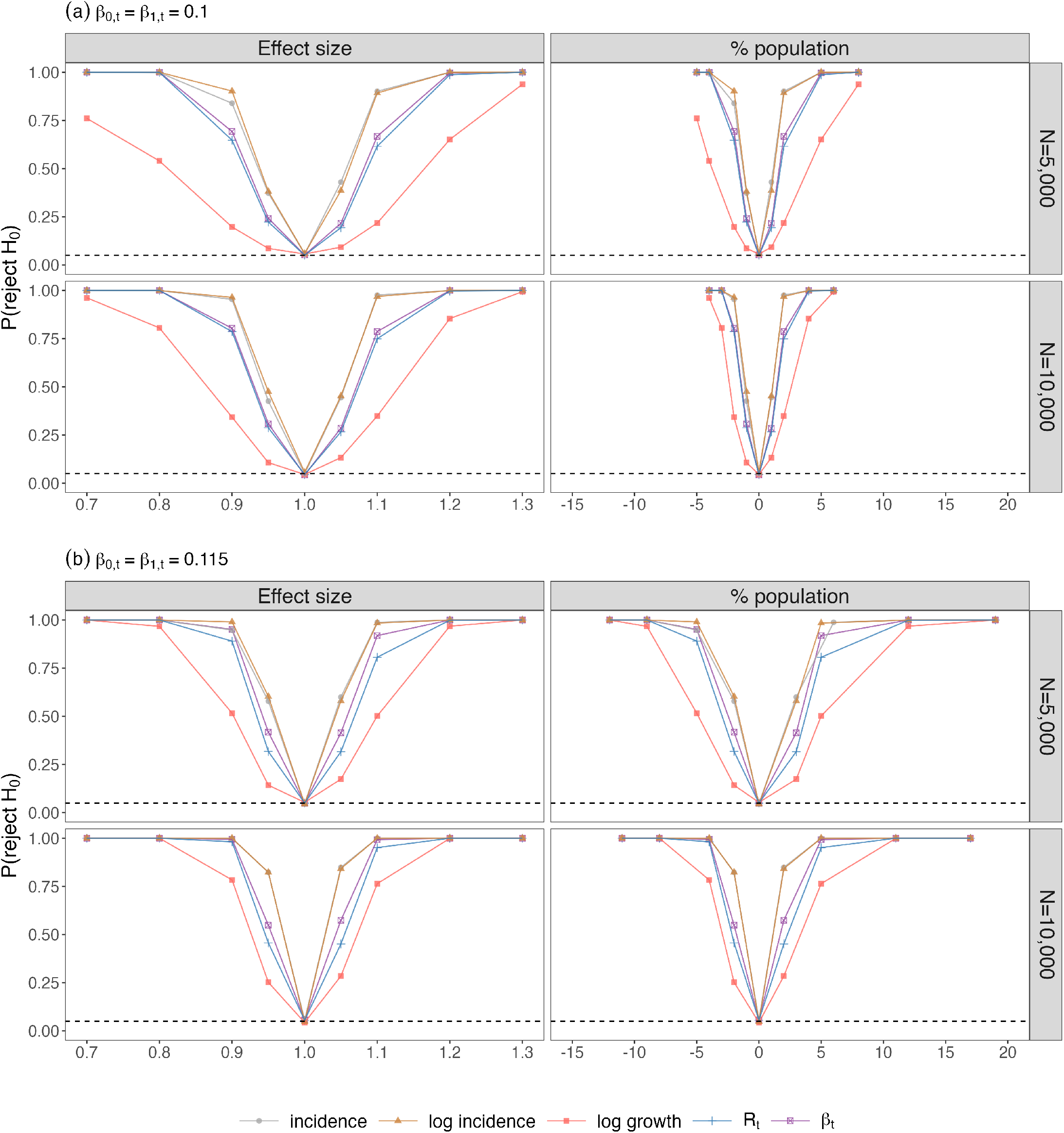
Power of models with all parameters equal between treatment and comparison groups. The top and bottom plots differ in the baseline effective contact rates: (a) *β*_0,*t*_ = *β*_1,*t*_ = 0.1, and (b) *β*_0,*t*_ = *β*_1,*t*_ = 0.115. Within each plot, left and right panels differ in the scale of effect size: the left plots the effect size as a multiplicative effect on baseline *β*_*d*,*t*_, and the right the percentage change in cumulative incidence. Across rows, we vary the population size. We generated data from an SIR model per Eq. 1. Solid colored lines indicate the average probability of rejecting the null over 1000 simulations. The dashed line indicates a significance level of 0.05.

In this case, all specifications controlled type I error (horizontal dashed line at 0.05) absent a treatment effect. With a non-null treatment effect, incidence and log incidence specifications produced the highest power, with minimal difference between the two. Of the remaining specifications, the log growth specification had the lowest power, while log *R*_*t*_ and log *β*_*t*_ had greater power, albeit slightly less than incidence and log incidence specifications. For example, with a treatment effect corresponding to a 5% increase in cumulative incidence over 6 weeks (e.g., 500 additional infections per a population of 10,000 individuals), the log growth specification had 80% power, while the other four model specifications had power exceeding 95%.

When the baseline effective contact rate differs between treatment and comparison groups, all methods except log *β*_*t*_ produced biased treatment effect estimates, failing to control type I error at *δ* = 1 (Figure 3). Although the fraction of the susceptible population depleted was lower with *N* = 10, 000, type I error remained higher than in the *N* = 5, 000 due to higher power with a larger population.

**Fig. 3.**
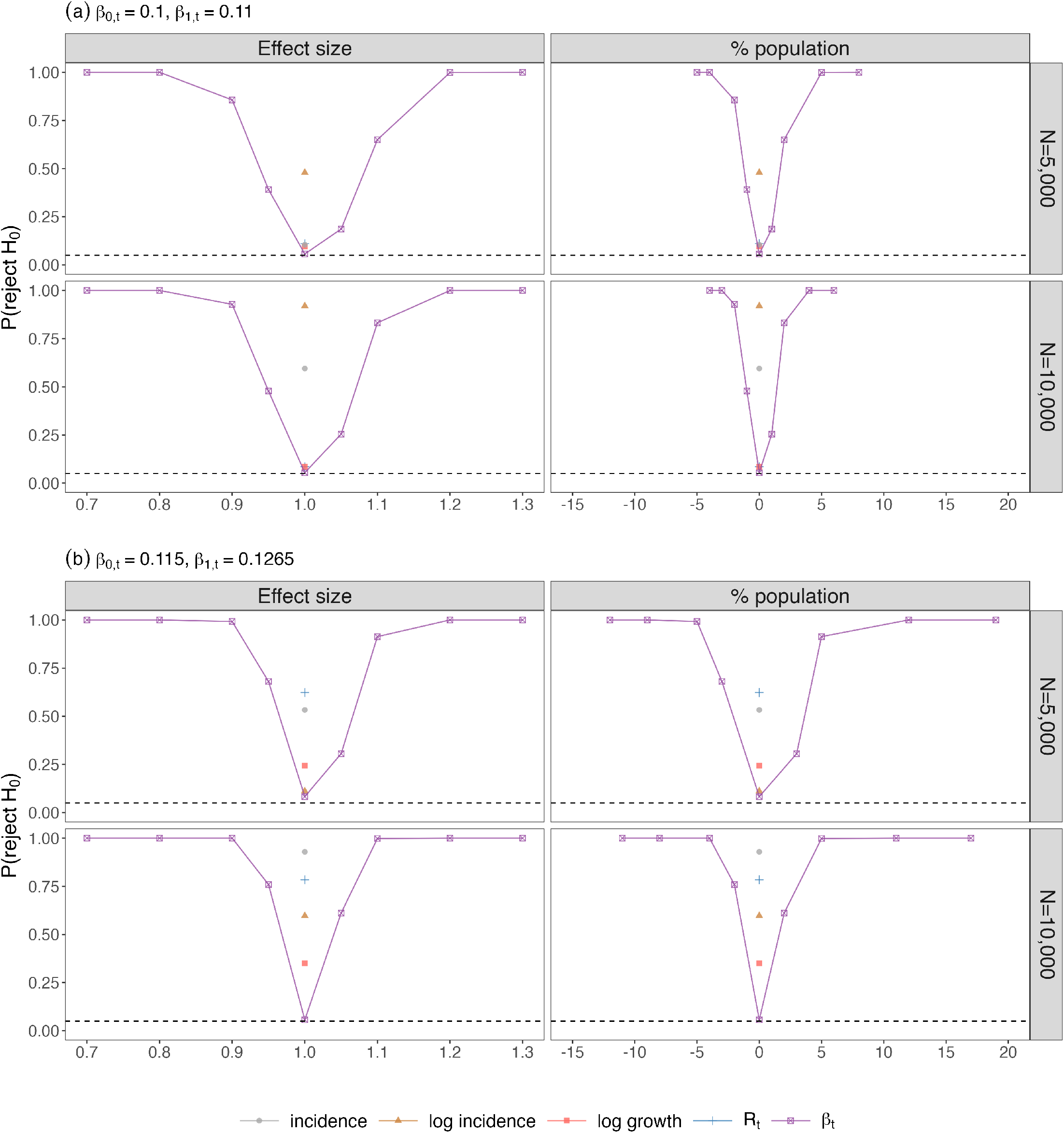
Power of models with baseline effective contact rate in treated units to be 10% greater than that of untreated units. The top and bottom plots differ in the scale of effect size: (a) *β*_0,*t*_ = 0.1, *β*_1,*t*_ = 0.11, and (b) *β*_0,*t*_ = 0.115, *β*_1,*t*_ = 0.1265. The left and right panels differ in the scale of effect size: the left plots the effect size as a multiplicative effect on baseline *β*_*d*,*t*_, and the right the percentage change in cumulative incidence. Across rows, we vary the population size. We generated data from an SIR model per Eq. 1. The solid colored line indicates the average probability of rejecting the null over 1000 simulations in the *β*_*t*_ model. Points at *δ* = 1 indicate the failure to control type I errors in all other models. The dashed line indicates a significance level of 0.05.

## Case Studies

### Removing school mask mandates in Massachusetts

#### Background

We last explore the implications of different specifications through two published case studies. We first consider an analysis of school mask mandates. On February 28, 2022, Massachusetts lifted its universal mask requirement in schools. Seventy of the 72 school districts lifted mask mandates shortly thereafter, but two districts, Boston and Chelsea, did not remove mask requirements until June 2022. The *New England Journal of Medicine* published a staggered DiD analysis estimating the impact of district-level mask policies, with weekly COVID-19 cases per 1000 as the outcome variable, using data from September 2021 to June 2022 (5). The analysis showed a statistically and practically significant increase in cases (an additional 45 cases per 1000 students and staff) associated with the removal of school mask mandates. We extended this analysis with log incidence and log growth specifications. We did not estimate effective reproduction number and effective contact rate estimates because the these are not typically estimated for subpopulations or below the county-level (22, 31).

#### Re-analysis methods

We applied the authors’ DiD approach to incidence, log incidence, and log growth models, which is similar to the specification in **Estimation**, but accounts for staggered treatment roll-out by estimating and averaging group-time average treatment effects (*AT T* (*g, t*)), where groups are defined based on time of treatment. We first replicated their specification using the Callaway & Sant’Anna estimator (26), with results presented in Supplement Table A2. As log incidence and log growth specifications do not allow zero outcomes, we implemented these with Poisson regression, first using the Callaway & Sant’Anna estimator and then a similar alternative by Sun and Abraham (27), that more straightforwardly supports inference with Poisson regression. For both, we present the average over all *AT T* (*g, t*). We then calculated AMEs for log incidence and log growth treatment effects.

#### Re-analysis results

Figure A1 plots district-level COVID-19 cases per 1000 over the study period. Using the Callaway & Sant’Anna estimator, we replicated the authors’ original findings, with an estimated 46 additional cases per 1000 students and staff (Figure A2). With the Sun & Abraham estimator, there was similarly an increase of 48 cases per 1000 students and staff (95% CI: 40, 57) (Table 2).

**Table 2.** Results from re-analyzing the effect of face mask policies in empirical examples.

| Study | Length of post-intervention periods | Outcome specification | Treatment effect<sup>1</sup><br>(95% CI) | Average marginal effect<sup>2</sup> (95% CI) |
| --- | --- | --- | --- | --- |
| Removing school mask mandates in Massachusetts <sup>4</sup> | 5 weeks | Incidence | 8.6* (5.7, 11.5) | 8.6* (5.7, 11.5) |
|  |  | Log incidence | 1.62* (1.26, 2.09) | 6.9* (1.8, 10.8) |
|  |  | Log growth | 0.97 (0.77, 1.22) | 8.6 (−10.9, 16.6) |
|  | 15 weeks | Incidence | 48.1* (38.9, 57.1) | 48.1* (38.9, 57.1) |
|  |  | Log incidence | 1.19 (0.90, 1.57) | 13.0 (−31.3, 47.1) |
|  |  | Log growth | 0.89 (0.70, 1.13) | −71.4 (−3036.0, 120.5) |
| Mask mandates in Kansas counties <sup>5</sup> | 19 weeks | Incidence | −21.6* (−28.3, −14.8) | −21.6* (−28.3, −14.8) |
|  |  | Log incidence | 0.33* (0.22, 0.59) | −55.1* (−101.2, −19.1) |
|  |  | Log growth | 0.96 (0.74, 1.25) | −9.5 (−2139.8, 25.3) |
| | | Log $R_t$ | 0.97 (0.90, 1.05) | −6.1 (−22.2, 8.3) |
| | | Log $\beta_t$ | 0.95<sup>†</sup> (0.86, 1.01) | −10.8<sup>†</sup> (−36.0, 2.6) |

<sup>1</sup> Treatment effect refers to the point estimate in coefficients obtained from OLS (incidence model) or average relative risk per unit from Poisson regression (other four models)
<sup>2</sup> Average treatment effects are imputed as described in Section **Average marginal effects** and Appendix E. AMEs for log $R_t$ and log $\beta_t$ models are calculated assuming an SEIR transmission process per Section F
\* represents significance at $\alpha = 0.05$
<sup>†</sup> represents significance at $\alpha = 0.10$

Figure A1 suggests that clusters do not have the same initial levels. Over the full study period, estimated treatment effects declined with a log incidence specification to 13 (95% CI: -31, 47) cases per 1000 students and staff (Table 2). The point estimate in the log growth specification was near 0, but highly uncertain; we could not rule out differences up to 120 cases per 1000 students and staff. However, with a shorter post-intervention horizon, both the incidence and the log incidence specifications detected a statistically and practically significant increase of 7-9 cases per 1000 students and staff over 5 weeks following the lifting of school mask mandates, with an effect of similar magnitude (but more uncertainty) in the log growth specification.

### Kansas mask mandates

#### Background

On July 3, 2020, Kansas adopted an executive order requiring face masks in most indoor and outdoor public places. As this was initially only adopted by 15 counties (32), *JAMA Network Open* published a DiD analysis evaluating the effect of county-level mask mandate adoption on daily COVID-19 rates (33). The authors used incidence specifications, where the 7-day moving average of COVID-19 cases per 100,000 was used as the outcome variable, and the policy was evaluated at 21 days after the executive order was effective (i.e., July 24, 2020 was the intervention time) to account gradual uptake and the lagged impact of behavior on cases. The authors found a statistically and practically significant reduction in COVID-19 cases (-20 cases per 100,000 with 95% CI: -27 to -14) following the executive order.

#### Re-analysis methods

We first replicated the authors’ original specification with small modifications:

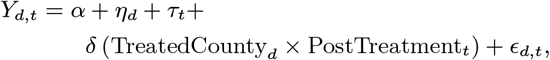

where the outcome *Y*_*d*,*t*_ was the county-level number of cases per 100,000 population (i.e., an incidence model), and the remaining variables and coefficients were defined as in **Estimation**. In our re-analysis, we aggregated data to the week-level and excluded counties with zero reported COVID-19 cases prior to the announcement of mask mandates. We also excluded covariates (an indicator for no reported cases and the number of days since the first recorded case in the county) to avoid recent methodological concerns about verifying the parallel trends assumption conditional on covariates (34).

We then re-analyzed the Kansas mask mandate example with the other four specifications. For log *R*_*t*_ and log *β*_*t*_ models, we obtained county-level data on *R*_*t*_ from COVIDestim nowcasting (31) and translated this to *β*_*t*_ by dividing *R*_*t*_ by the estimated susceptible fraction as illustrated in Proposition 6. We used estimated the susceptible fraction using cumulative infections from COVIDestim nowcasting (31) and each county’s 2019 population data from the Census Bureau (35) to impute susceptible fractions. For AMEs with log(*R*_*t*_) and log(*β*_*t*_), we used an SEIR model, parameterizing for COVID-19, with an average of 5 days of infectiousness and a 3-day incubation period (36).

#### Re-analysis results

Our treatment effect estimate using a slightly modified incidence model was − 22 cases per 100,000 (95% CI: − 28 to − 15), very similar to the original estimate (-20 cases per 100,000 with 95% CI: -27 to -14).

Table 2 summarizes the estimated treatment effects and average marginal effects obtained using all five specifications. We observed a similar treatment effect in the log incidence specification compared to the incidence specification. Effects were smaller and insignificant for log growth and log *R*_*t*_ specifications. However, in this application, many counties experienced considerable susceptible depletion (e.g., the estimated susceptible fraction dropped by 40% in Nemaha and 43% in Norton over the study period), a potential threat to the validity of log growth and log *R*_*t*_ models. By contrast, the log *β*_*t*_ specification produced a significant treatment effect at 90% confidence level, with an average marginal effect of -11 cases per 100,000.

## Discussion

DiD is a popular method for observational causal inference in health policy. This arises in part from its flexibility: DiD allows researchers to estimate treatment effects, even absent comparison groups that exactly match the treatment group. DiD is traditionally agnostic as to why treatment and comparison groups differ in level but are believed to have matching trends (11). However, by synthesizing DiD with a mechanistic model for infectious disease transmission, we showed how context-specific theory informs interpretation of the parallel trends assumption.

Assuming an underlying SIR framework, our paper first formalized the epidemiological conditions under which the parallel trends assumption holds for the three common model specifications in the COVID-19 literature: incidence, log incidence, and log growth. Although modeling incidence was popular in the COVID-19 medical and health policy literature (Appendix A), this approach requires identical data-generating processes between all treated and comparison units, as others have previously noted (4). Modeling log incidence allows for different numbers of initial infections but still requires equal effective contact rates and an “infinite susceptible population” assumption. Nevertheless, because we found incidence and log incidence specifications to have very similar power, we recommend that researchers default to the latter, binning data or applying Poisson regression if there are zero-valued outcomes (37). By contrast, although log growth specifications allow for more flexibility, they comes with a significant power cost.

We argued that the log growth specification approximates assuming parallel trends in the effective contact rate (e.g., assuming similar behavior but different vaccination rates across units). Both modeling log growth and the log of effective reproduction number (a similar but higher power alternative) may produce biased estimates in the context of susceptible depletion. Our most robust alternative is therefore modeling the log of effective contact rate (*β*_*t*_) directly. We showed that this approach has higher power than modeling log growth, performs well under more flexible assumptions, and can be estimated with zero-valued outcomes using Poisson regression. We also demonstrated that this can be applied to more diverse and complex underlying transmission processes. As a result, our work provides a useful bridge between post-hoc policy evaluation and prospective transmission dynamic models that serve as a backbone of infectious disease epidemiology.

Last, we apply different model specifications to previously published examples, highlighting trade-offs between bias and power and emphasizing the importance of evaluating and discussing the plausibility of assumptions required for a chosen specification.

This work has several limitations. First, even our most robust specifications may not be plausible in practice. Nevertheless, our approach allows for more flexibility than many proposed alternatives. For example, Callaway and Li (4) proposed an alternative estimator in the context of early COVID-19 that assumed unconfoundedness after conditioning on all pre-treatment period outcomes. This method performs well in the early stages of an epidemic when conditions across units in both groups are similar. However, it may be challenging to find exact matching units over the full epidemic history, especially when the effective contact rate is time-varying due to shifting precautionary behaviors or vaccination rates. Relatedly, although our approach can be extended to a wide array of models, we do not consider the implications of spillovers, which may obscure treatment effects.

Second, estimating the effective reproduction number and the effective contact rate can be challenging in practice. Many past studies (e.g., O’Driscoll et al. (20), Gostic (22)) have discussed challenges and proposed approaches for translating observed cases and deaths into estimates of infections and the effective reproduction number, which would be valuable when applying our methods. Last, our results highlight that in many cases, policy evaluations may be underpowered to detect effects of substantive interest. Future efforts could take a decision-analytic approach to integrate the results of policy evaluations with costs and benefits of implementing different model specifications.

Overall, our work provides a framework for integrating transmission dynamics into DiD and proposes novel methods to support rigorous evaluation of infectious disease policies.

## Data Availability

All data produced are available online and from previously published work

## ACKNOWLEDGMENTS

The authors gratefully acknowledge feedback from Tori Cowger, Jeremy Goldwasser, Kathryn T. Hall, Joseph Hogan, Youjin Lee, Eleanor Murray, Jonathan Roth, and Pedro Sant’Anna. We also greatly appreciate replication code shared by Donna Ginther. This work was supported by the Centers for Disease Control and Prevention through the Council of State and Territorial Epidemiologists (NU38OT000297-02).

## A. Literature Review

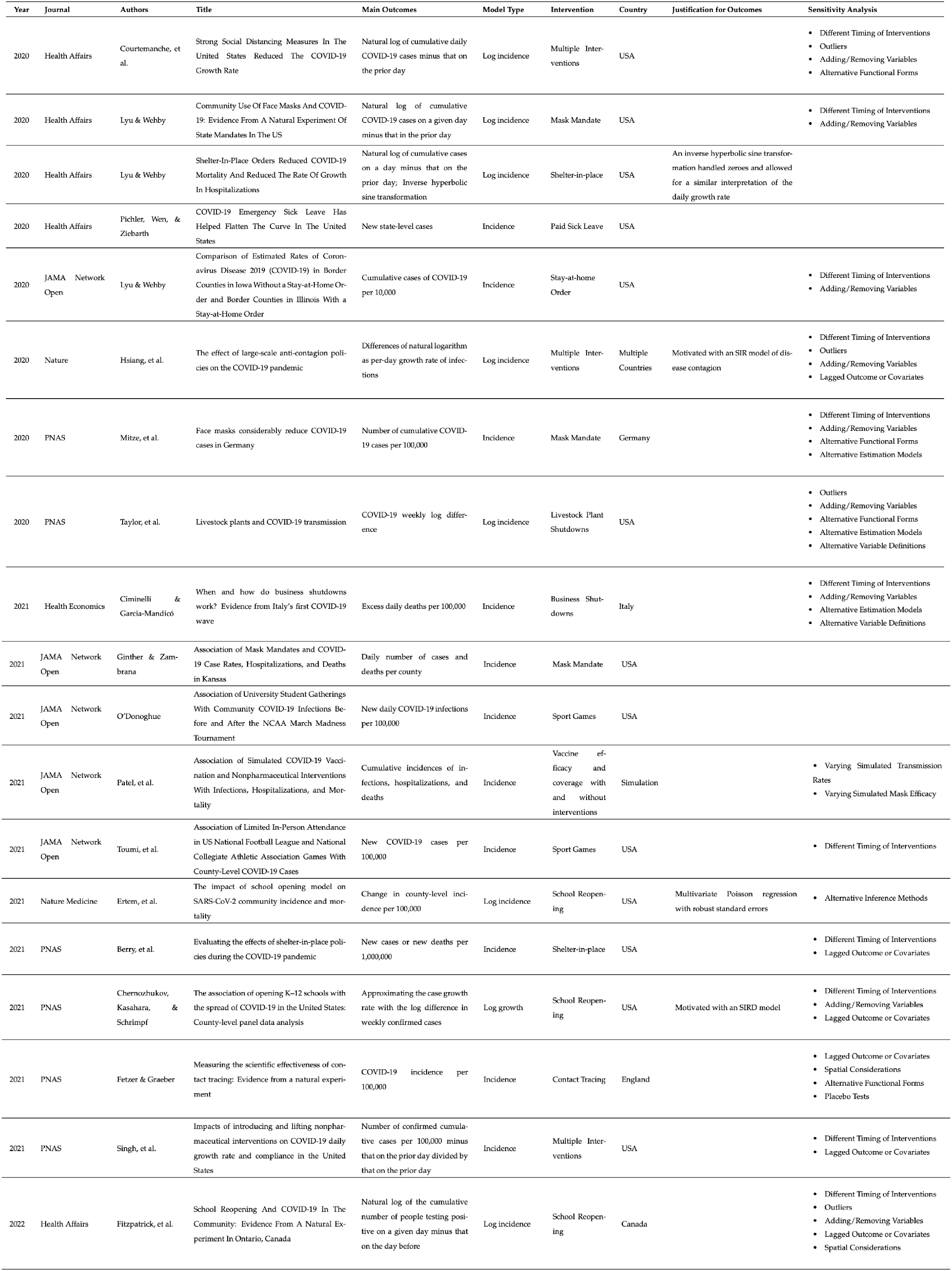
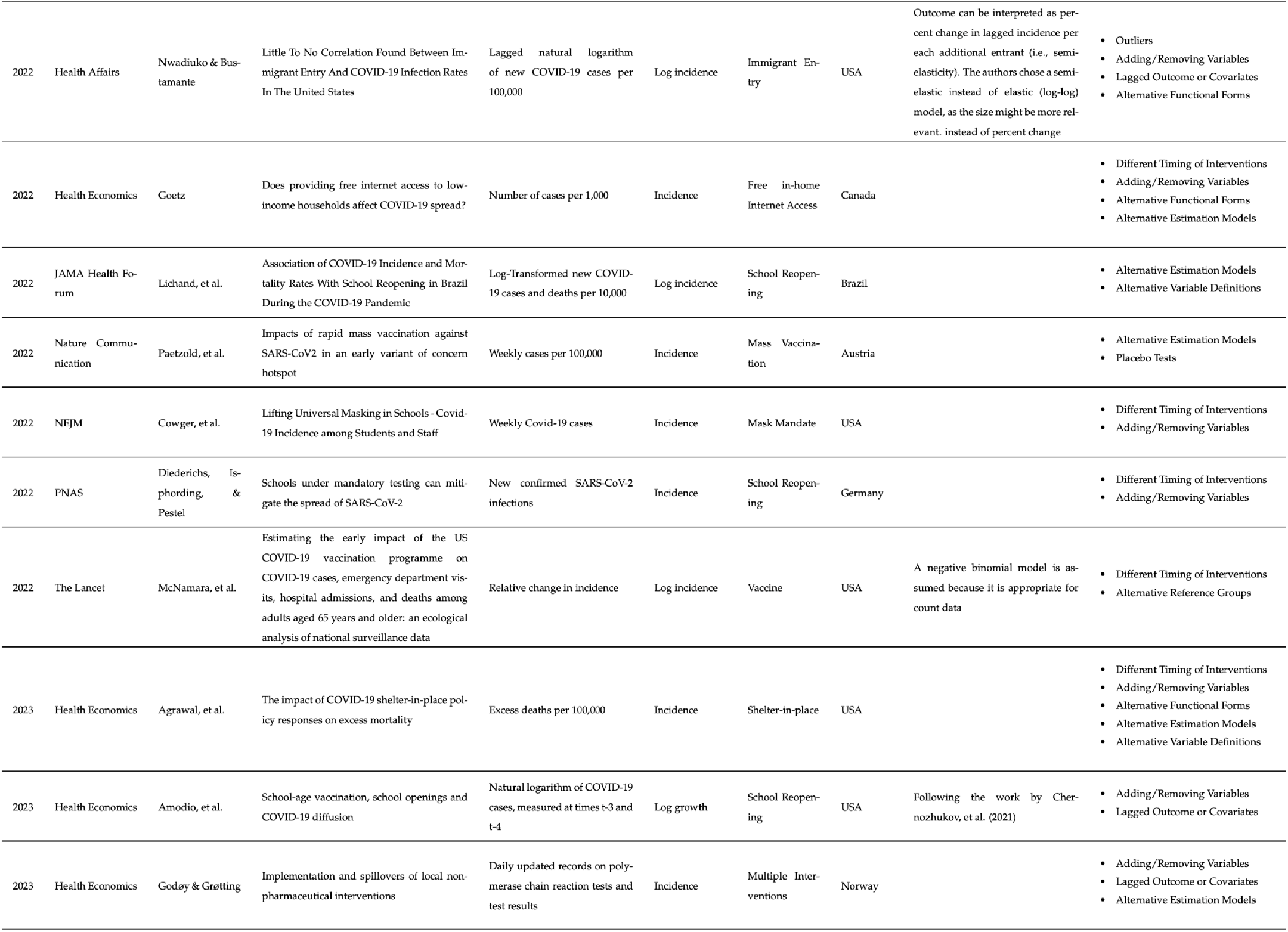

## B. Proofs

### Proposition 1

(Expected incidence). *Assuming an SIR data-generating process (Eq. 1) with initial conditions* {*S*_*d*,0_, *I*_*d*,0_, *R*_*d*,0_}, *expected incidence at time t* + 1 *can be written as:*

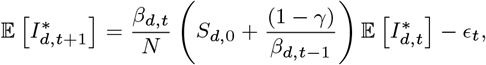

*where* 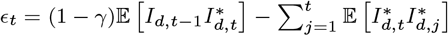.

*Proof*. Given {*I*_*d*,0_, *S*_*d*,0_, *R*_*d*,0_}, constant generation interval 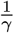, and effective contact rates (*β*_*d*,*t*_), we have per Eq. 1:

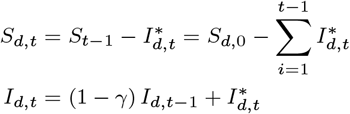

We denote incidence at time *t* + 1:

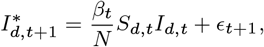

where 𝔼 [*ϵ*_*t*_] = 0 because 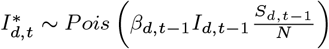.

Taking the expectation of both sides:

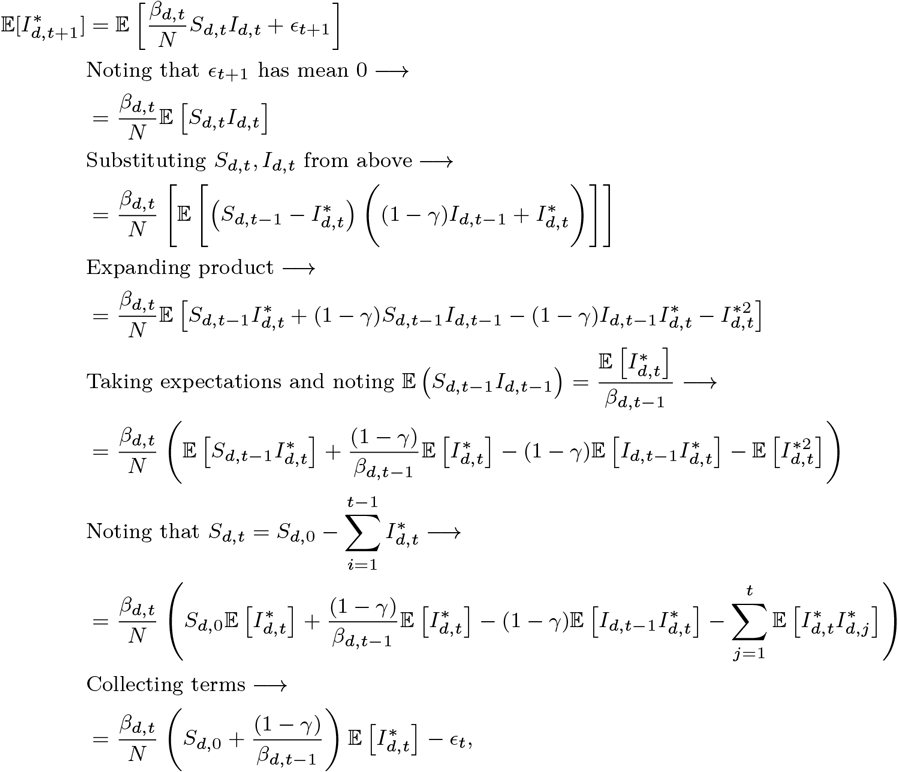

where 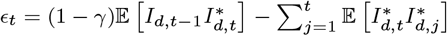.

### Proposition 2

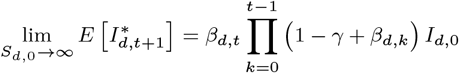

*Proof*. We prove Proposition 2 by induction. For explication, we write 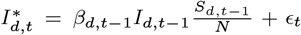, where 𝔼 [*ϵ*_*t*_] = 0 because 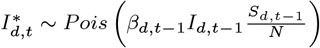. Population size *N* is assumed constant (Eq. 1) over time, with *N* = *S*_*d*,0_ + *I*_*d*,0_ + *R*_*d*,0_ for all *t*.

*Base case* (*t* = 1):

We first take the expectation of *I*_*d*,1_:

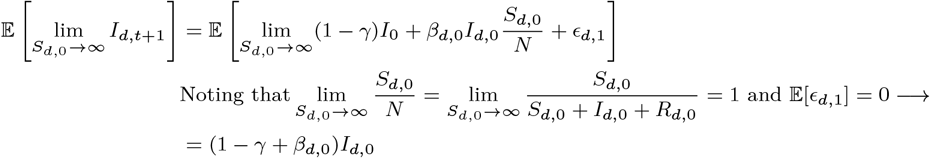

We then take the expectation of incidence at time *t* + 1 = 2:

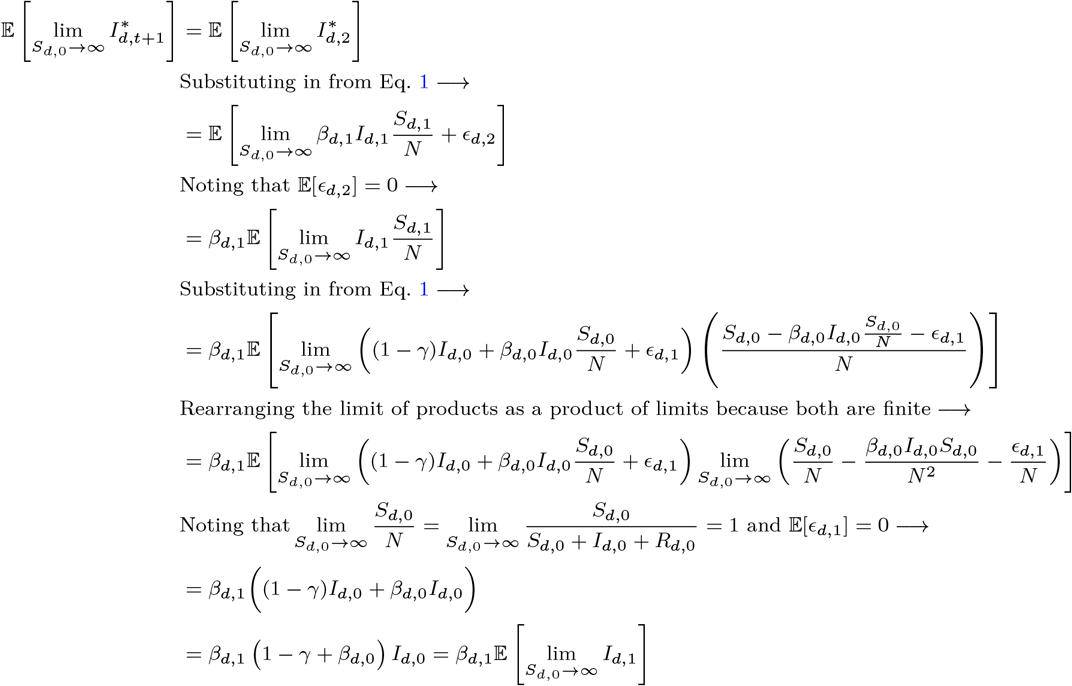

*Induction step:* Assume that:

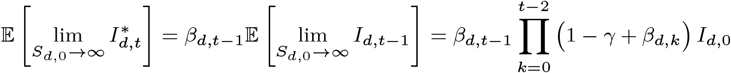

We will show that this implies that the same holds for the next time step, i.e.,

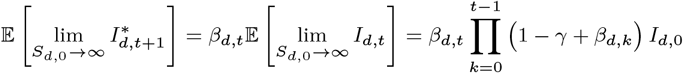

We have:

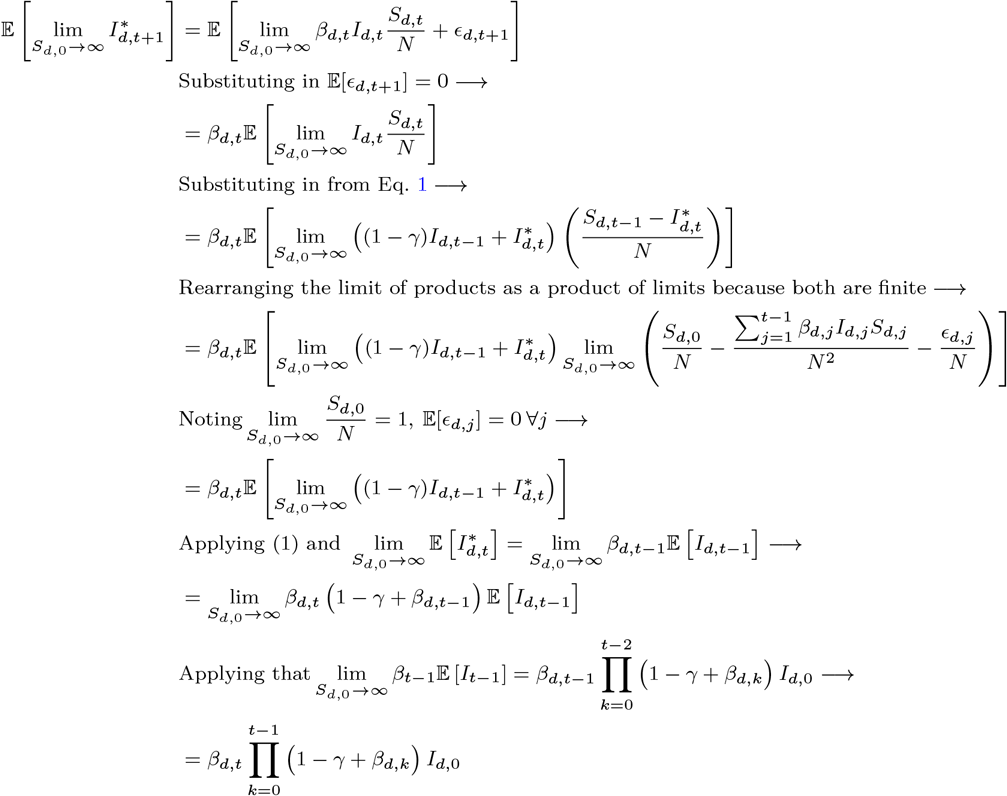

This completes the induction step.

Last, note that the dominated convergence theorem allows us to exchange limit and expectation because for all *S*_*d*,0_, 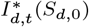 is stochastically dominated by 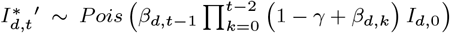 (i.e., 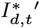 from in a modified SIR process in which 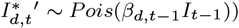 over all *d, t*. □

### Proposition 3

(Parallel trends: Incidence (infinite susceptible population)). *Assuming an SIR data-generating process (Eq. 1) and an incidence model specification* 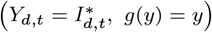, *the “infinite susceptible population” parallel trends assumption (Eq. 3) holds between t*_1_ *and t*_2_ *under the following conditions:*

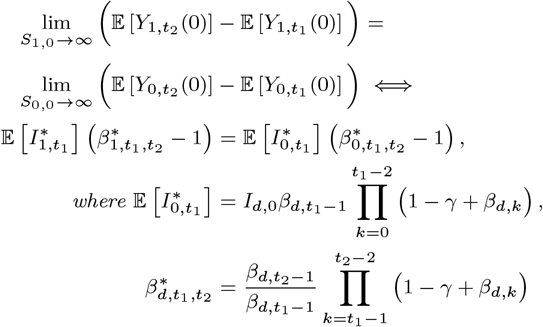

*Proof*. For *t*_1_, *t*_2_ ≥ 2 and *d* ∈ {0, 1}, we have:

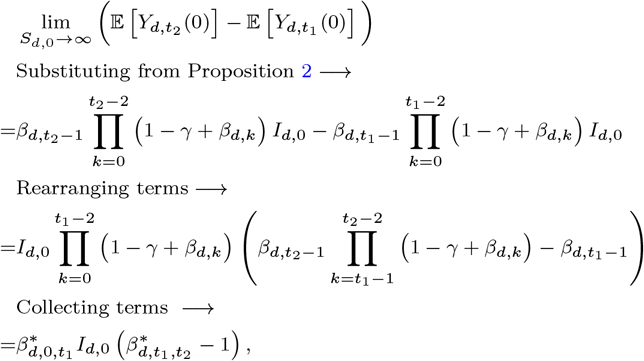

where 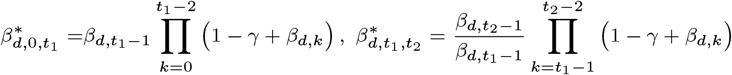

Substituting the above expression into the parallel trends condition, we obtain ^‖^:

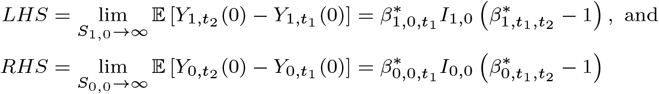

□

### Proposition 4

(Parallel trends: Log incidence (infinite susceptible population)). *Assuming an SIR data-generating process (Eq. 1) and a log incidence model specification* 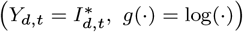, *the “infinite susceptible population” parallel trends assumption (Eq. 3) holds between t*_1_ *and t*_2_ *under the following conditions:*

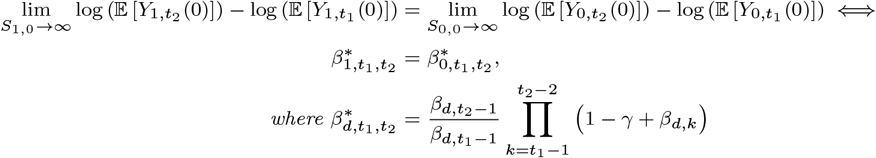

*Proof*. For *t*_1_, *t*_2_ ≥ 2 and *d* ∈ {0, 1}, we expand as follows:

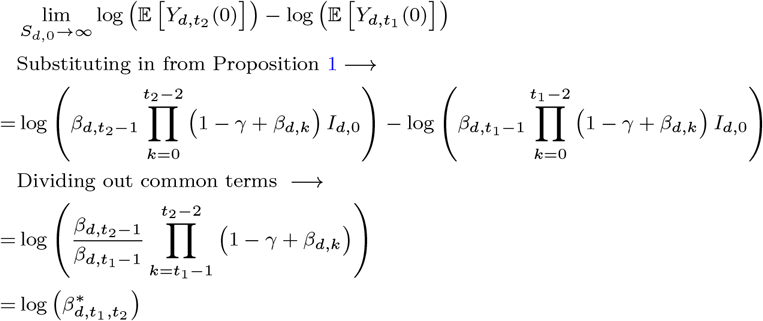

Therefore, the “infinite susceptible population” parallel trends assumption (Eq. 3) holds if and only if

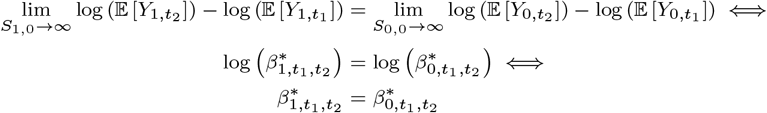

□

### Proposition 5

(Parallel trends: Log growth (infinite susceptible population)). *Assuming an SIR data-generating process (Eq. 1) and a log growth model specification* 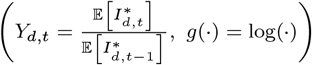, *the “infinite susceptible population” parallel trends assumption (Eq. 3) holds between t*_1_ *and t*_2_ *under the following conditions:*

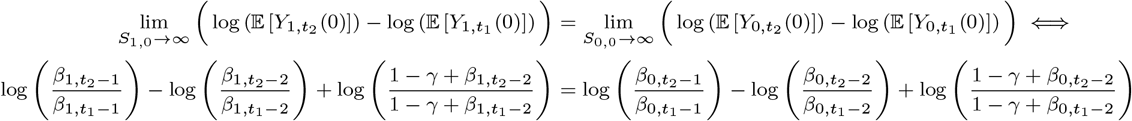

*Proof*. For *t*_1_, *t*_2_ ≥ 2 and *d* ∈ {0, 1}, we have:

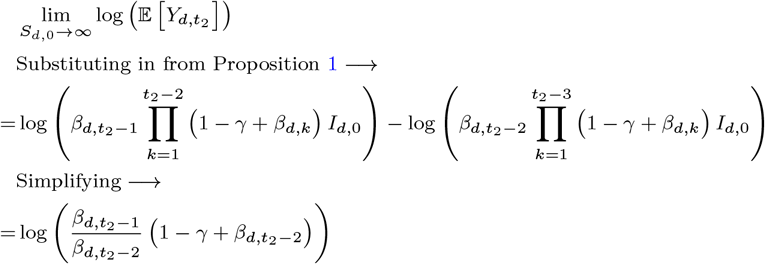

Similarly, 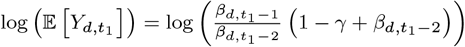. Therefore,

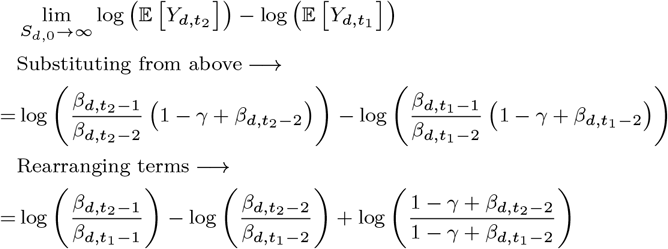

Substituting the above equation back to both sides of the parallel trends assumption defined in Eq. 3 completes the proof. □

### Proposition 6

(Cohort definition of *R*_*t*_). *Assume that the effective reproduction number is measured over a generation interval of length* 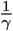 *for the cohort* 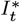 *becoming infectious at time t. We define the cohort effective reproduction number:*

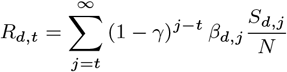

*Proof*. Eq. 1 defines the average number of secondary infections per infected individual at time *t* as 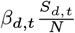. Of individuals who become infected at time , Eq. 1 also defines the fraction removed at each time step as 1− *γ*. This gives us the effective reproduction number corresponding to the cohort becoming infectious at time *t*:

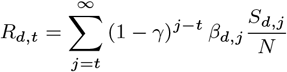

□

### Proposition 7

(Parallel trends: Log *R*_*t*_ (infinite susceptible population)). *Assuming an SIR data-generating process (Eq. 1), log-transformed effective reproduction number model specification (Y*_*d*,*t*_ = log (*R*_*d*,*t*_), *g*(·) = log(·)), *the “infinite susceptible population” parallel trends assumption (Eq. 3) holds for all t*_1_, *t*_2_ *if and only if*

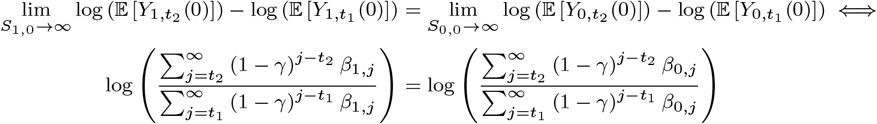

*Proof*. In Proposition 6, we defined 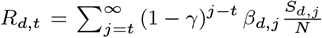. Substituting this formulation of *R*_*d*,*t*_ into the parallel trends assumption in Eq. 3, we obtain:

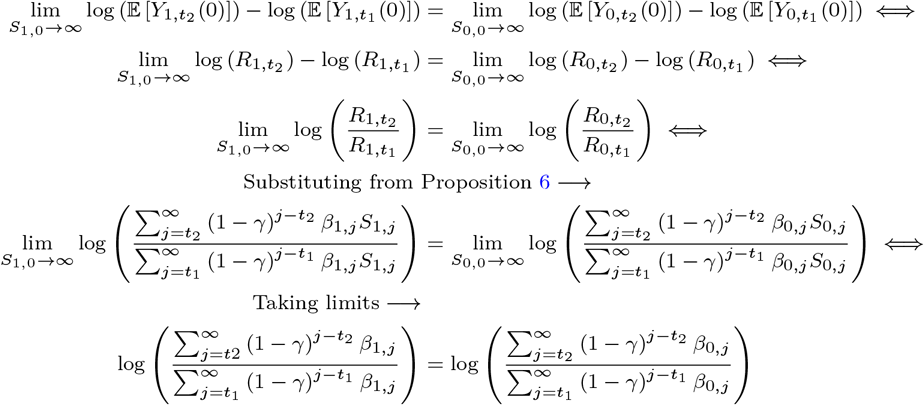

□

### Proposition 8

(Parallel trends: Log *β*_*t*_). *Assuming an SIR data-generating process and a log-transformed effective reproduction number specification (Y*_*d*,*t*_ = log (*β*_*d*,*t*_), *g*(·) = log(·)), *the parallel trends assumption holds for all t*_1_, *t*_2_ *> t* − 1 *if and only if*

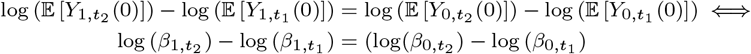

*Proof*. Proposition 8 follows from a direct substitution of the outcome *Y*_*d*,*t*_ = *β*_*d*,*t*_ into the log-transformed parallel trends assumption as defined in Eq. 3:

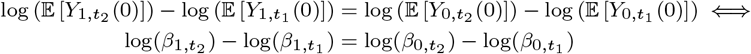

□

### Proposition 9

(Estimation of *β*_*d*,*t*_). *Assuming an SIR data-generating process (Eq. 1), with* 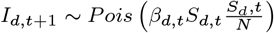, *the maximum likelihood estimator of β*_*d*,*t*_ *is:*

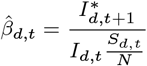

*Proof*. Because we assume:

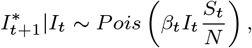

the likelihood (*L*) and log-likelihood (*𝓁*) functions can be defined:

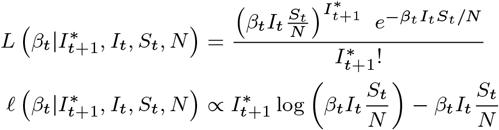

Setting 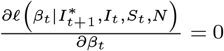 to obtain the maximum likelihood estimator:

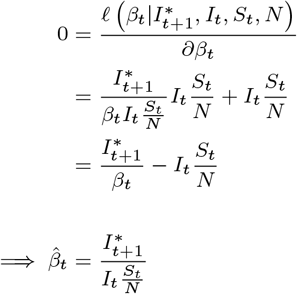

### Supplement Proposition 1

(Log transformation). *Assume that Eq. 3 holds with a log link, such that:*

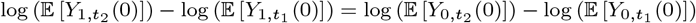

*Then,*

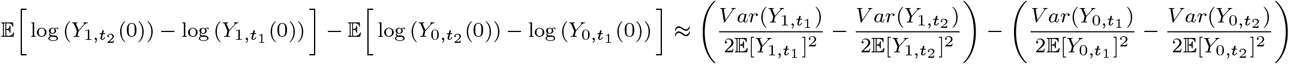

*Proof*. We can approximate 𝔼 [log(*Y*_*d*,*t*_)] with a second-order Taylor series expansion (38):

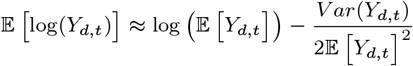

Substituting this approximation for each term in 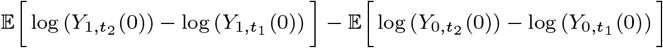 completes the proof. □

### Corollary 1.

*If the outcome is log incidence in Supplement Proposition 1*, 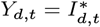 *with a log link, then*,

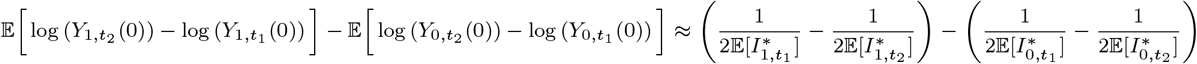

*Proof*. Per Eq. 1, we assume the incidence 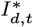 follows a Poisson distribution. As a result, the second-order term becomes:

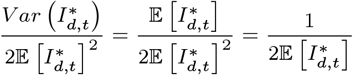

Substituting this into the result from Proposition 1 completes the proof. □

### Supplement Proposition 2

(DiD with time-step aggregation). *Assume that the parallel trends assumption (Eq. 3) holds with a log link for every pair of individual pre- and post-intervention time steps between the average outcome in the treated and comparison groups. That is, for any pre-intervention time step t*_1_ *and post-intervention time-step t*_2_, *we assume*

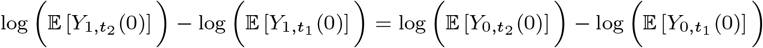

*Then, the aggregated parallel trends assumption (Eq. 5) also holds:*

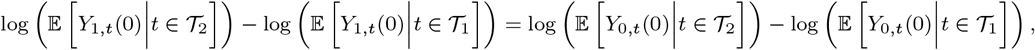

*where 𝒯*_1_ *and 𝒯*_2_ *denote aggregations of pre- and post-intervention time periods*.

*Proof*. By imposing parallel trends (Eq. 3) for any pair of individual pre- and post-intervention time steps, we have for any *t*_1_ and *t*_2_,

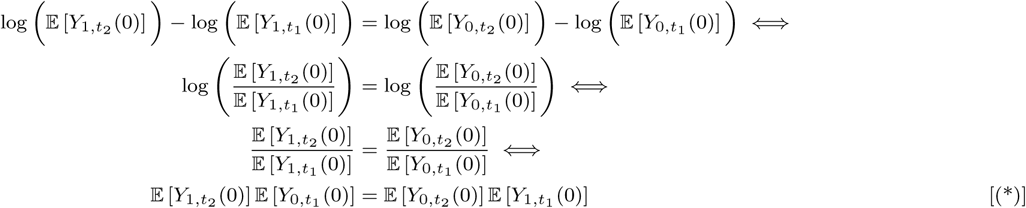

For the parallel trends condition to hold on the aggregated time periods *𝒯*_1_ and *𝒯*_2_, we want to show

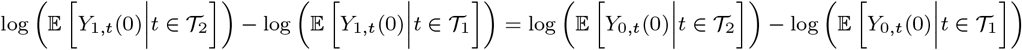

Note that if aggregation occurs by summing, rather than averaging, both the LHS and RHS are multiplied by the number of time steps, which does not affect the result. For each side of the above equation, the difference in log expected outcomes can be written as a sum of time steps:

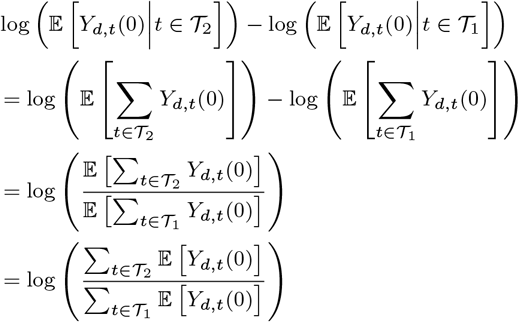

Therefore, Eq. 5 holds for *𝒯*_1_ and *𝒯*_2_) if and only if

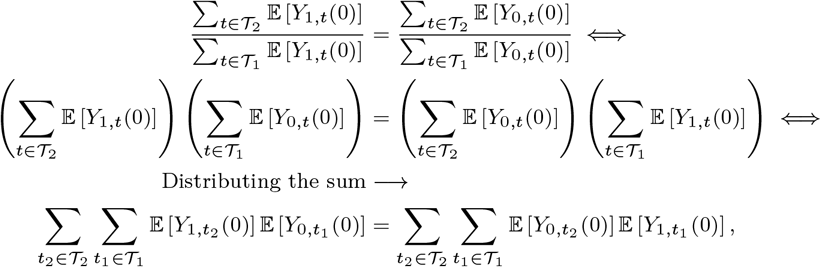

where the last equality is true because Eq. 3 holds for every pair of pre-intervention *t*_1_ and post-intervention *t*_2_.

### Corollary 2.

*Assume that the parallel trends assumption (Eq. 3) holds with a log link for every pair of individual pre- and post-intervention time steps between the average outcome in the treated and comparison groups as in Supplement Proposition 2. Then the extended parallel trends assumption (Eq 6) holds on the transformed average differences between pre- and post-treatment outcomes across treated and untreated groups:*

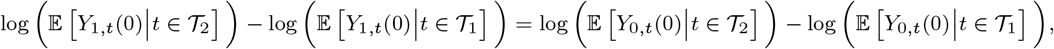

*where 𝒯*_1_ *and 𝒯*_2_ *are set of pre- and post-treatment time periods, respectively*.

*Proof*. The proof follows directly from Supplement Proposition 2, with only the definitions of *𝒯*_1_ and *𝒯*_2_ changed.

## C. Figures

**Fig. A1.**
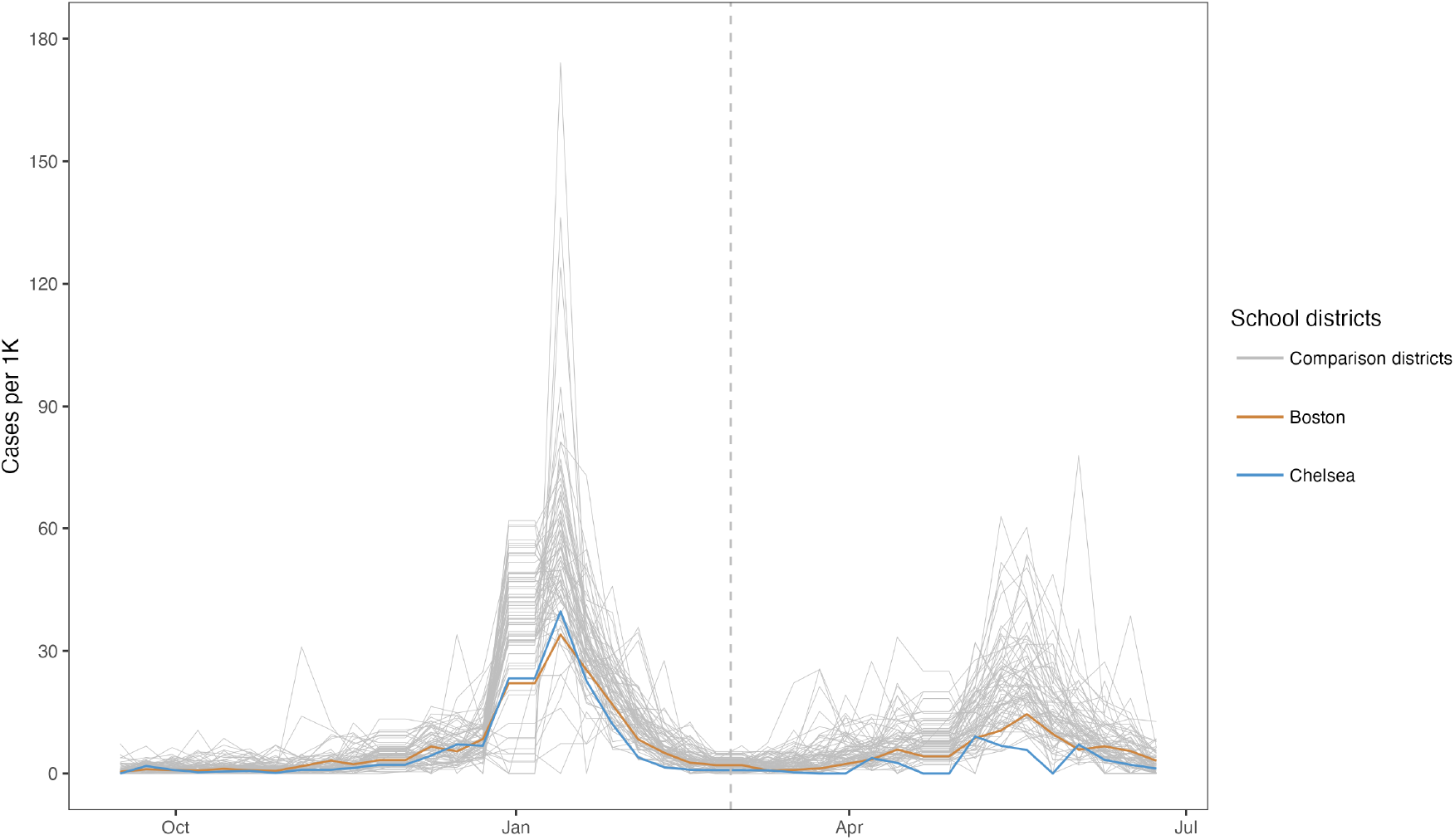
COVID-19 cases per 1000 students and staff in 72 school districts in Massachusetts. Each line plots the case trajectory in one school district. The dashed vertical line represents the time at which the state-level mask mandate was lifted.

**Fig. A2.**
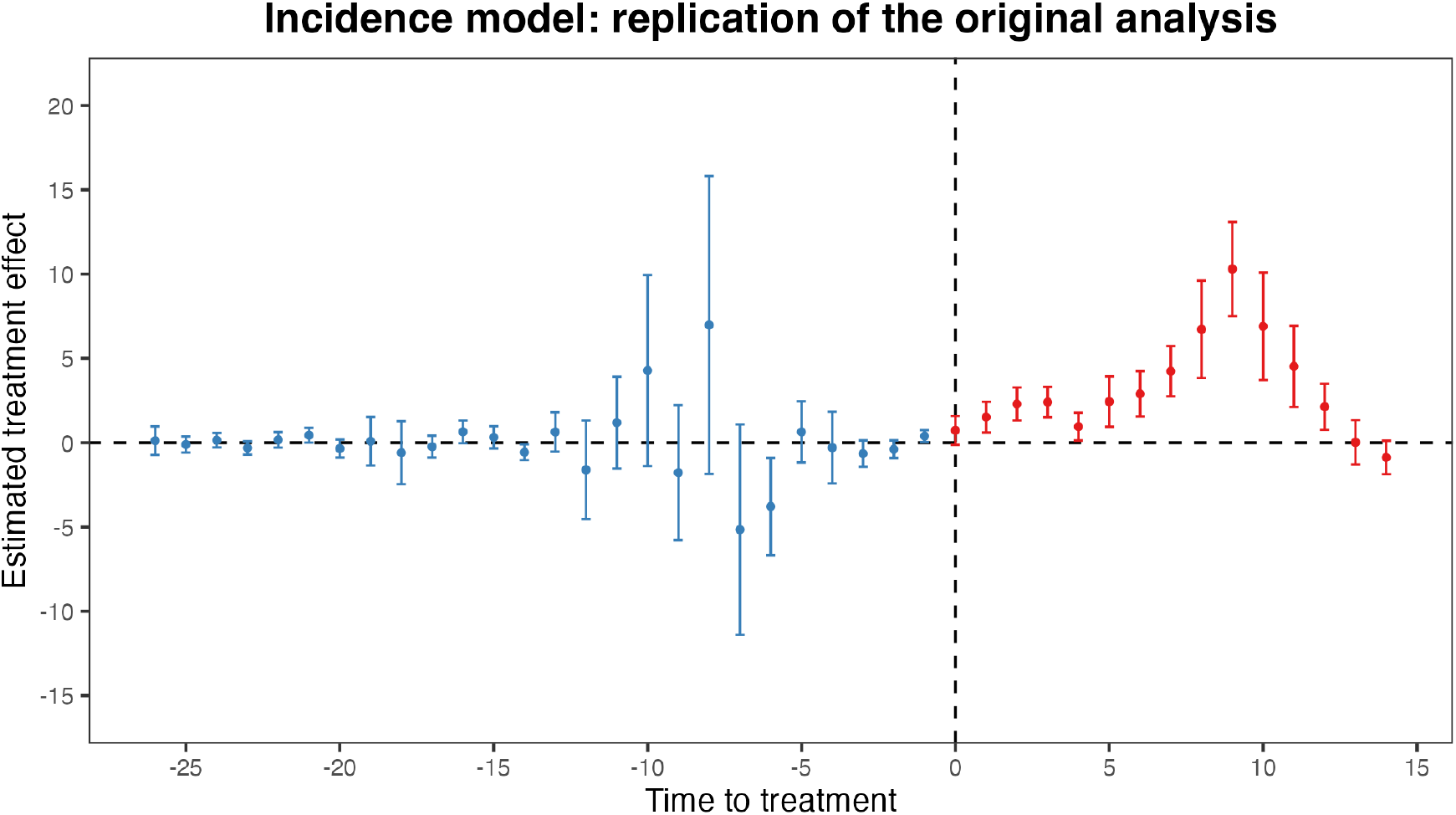
Event study plot from replicating the original analysis Cowger et al. (5) The estimated treatment effects with associated 95% confidence intervals obtained from fitting the incidence specification, which is the model specification in the original analysis, are plotted against the time to treatment.

**Fig. A3.**
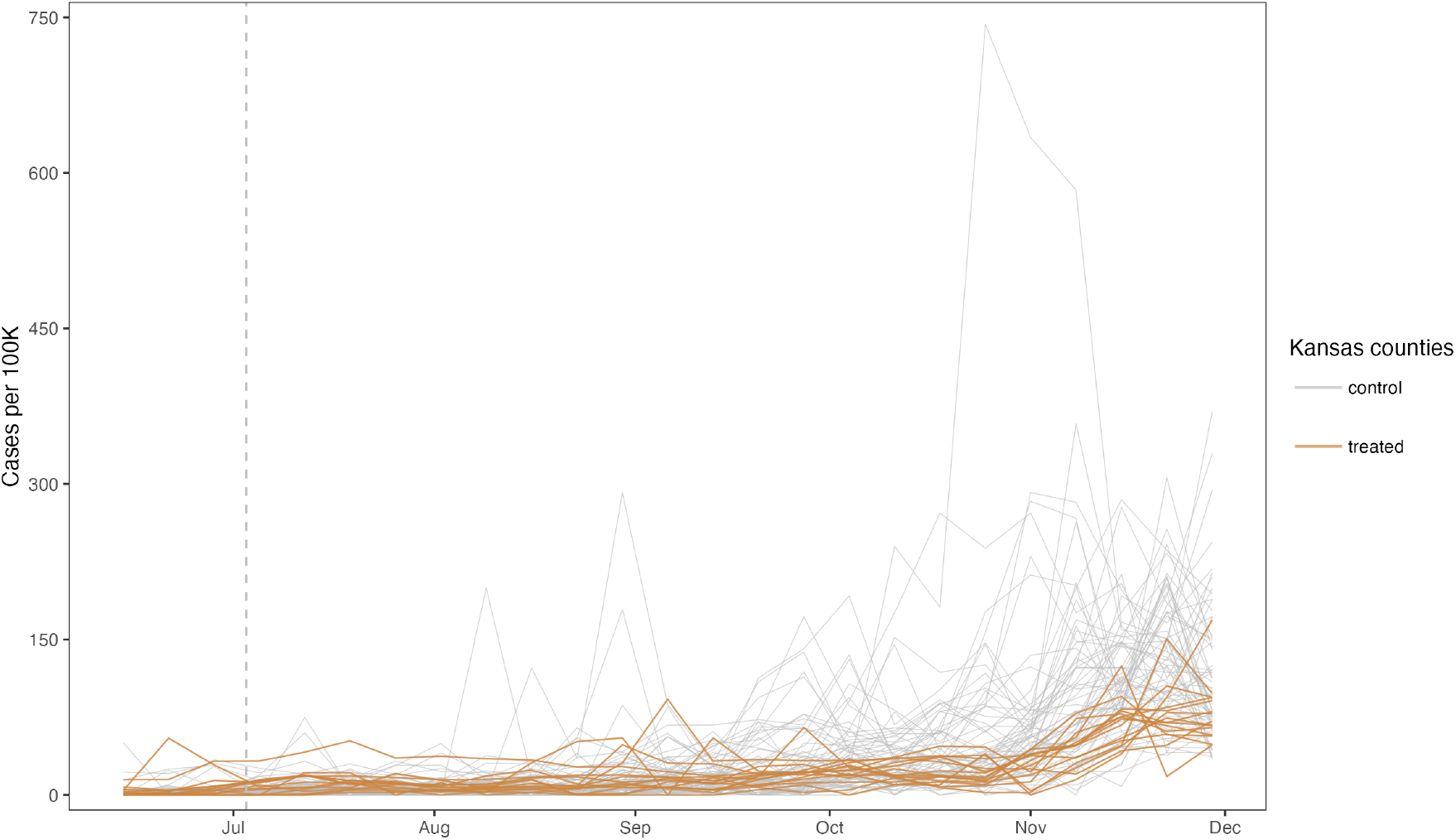
COVID-19 cases per 100,00 population in Kansas. Each line plots the case trajectory in one county in Kansas. The dashed vertical line represents the time at which the executive order took effect.

## D. Tables

**Table A1.**
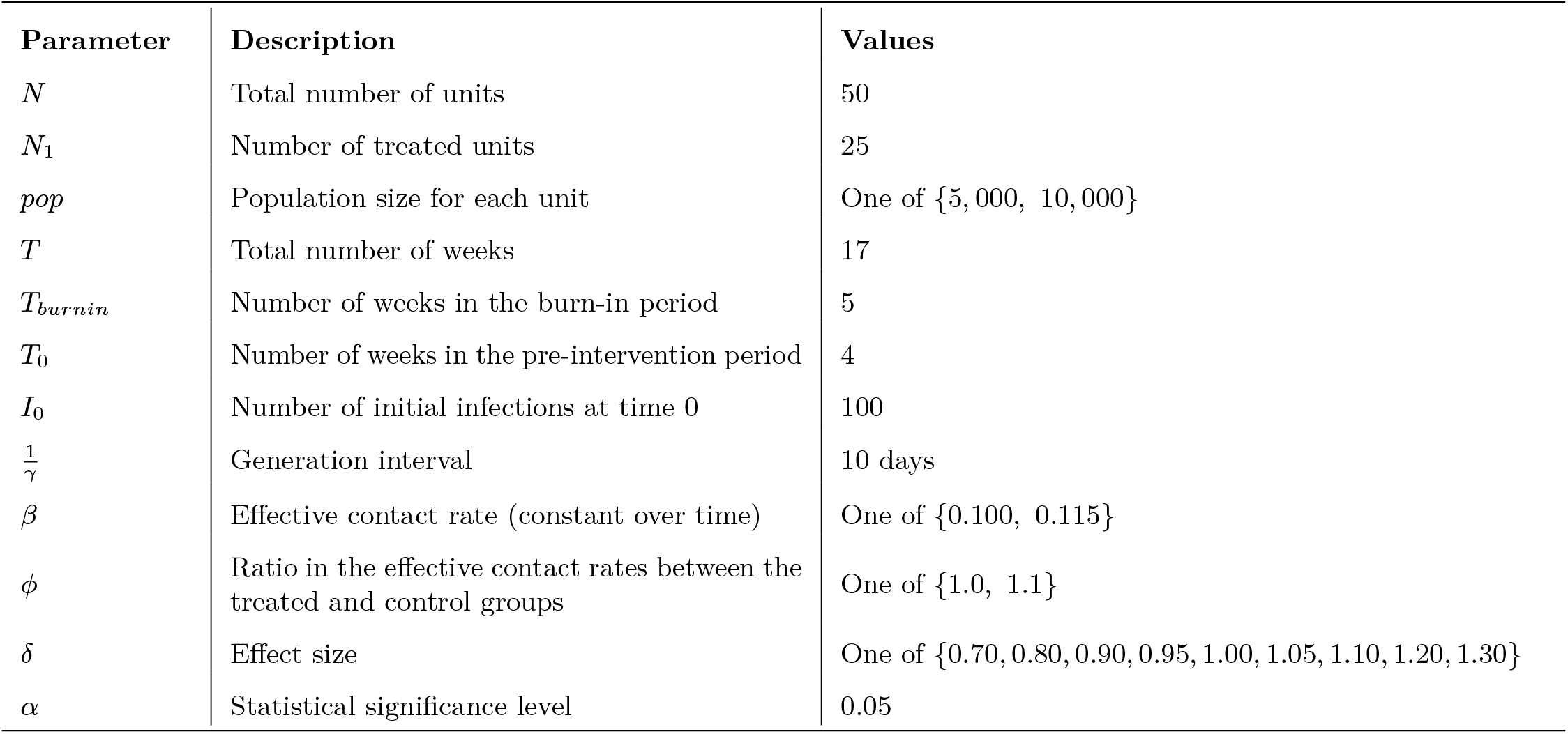
Simulation parameters.

| Parameter | Description | Values |
| --- | --- | --- |
| $N$ | Total number of units | 50 |
| $N_1$ | Number of treated units | 25 |
| $pop$ | Population size for each unit | One of {5,000, 10,000} |
| $T$ | Total number of weeks | 17 |
| $T_{burnin}$ | Number of weeks in the burn-in period | 5 |
| $T_0$ | Number of weeks in the pre-intervention period | 4 |
| $I_0$ | Number of initial infections at time 0 | 100 |
| $\frac{1}{\gamma}$ | Generation interval | 10 days |
| $\beta$ | Effective contact rate (constant over time) | One of {0.100, 0.115} |
| $\phi$ | Ratio in the effective contact rates between the treated and control groups | One of {1.0, 1.1} |
| $\delta$ | Effect size | One of {0.70, 0.80, 0.90, 0.95, 1.00, 1.05, 1.10, 1.20, 1.30} |
| $\alpha$ | Statistical significance level | 0.05 |

**Table A2.** Results from re-analyzing the effect of removing school mask mandates on COVID-19 cases in Massachusetts using Callaway and Sant’Anna estimator.

| Outcome specification | Treatment effect<sup>1</sup> | Average marginal effect<sup>2</sup> |
| --- | --- | --- |
| Incidence | 47.4 | 47.4 |
| Log incidence | 2.0 | 10.0 |
| Log growth | -2.2 | -179.5 |

<sup>1</sup> Treatment effect refers to the point estimate in coefficients obtained from OLS (incidence model) or Poisson regression (log incidence and log growth models) <sup>2</sup> Average treatment effects are imputed as described in Section **Average marginal effects** and Appendix E.

## E. Algorithms

### Algorithm 1

(Estimation of average marginal effects for log incidence and log growth specifications) Given the observed outcomes in the treated units, 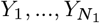, and the estimated ATT, 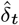, we impute the AME as follows:

1. Calculate the fitted untreated potential outcome for the treated group in the scale of the model specification for each treated unit *i* and post-intervention period *t > T*_0_ using the observed empirical outcome trajectory and the estimated 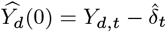
2. Recover the fitted untreated potential outcome for the treated unit *i* in the case scale, 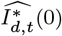), from *Ŷ*_*d*,*t*_(0) according to the definition of model specifications per Table 1. For log growth, we take the last period prior to intervention as baseline, and construct the untreated potential outcomes by dividing the baseline outcome by the fitted treatment effect coefficient. We repeat division for each post-intervention period to recover the untreated trajectories for the treated units.
3. Calculate the difference between the observed treated outcome and the fitted control potential outcome trajectories to obtain the marginal effect (ME) for each unit *i* over the entire post-intervention time periods: 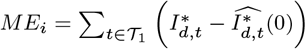
4. The AME is the average of the calculated differences over all treated units: 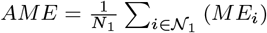

### Algorithm 2

(Estimation of average marginal effects for log *R*_*t*_ or log *β*_*t*_ models) For COVID-19, we assume on average 5 days of infectiousness and 3 days of mean exposure period (36). We use input data on the initial susceptible fraction and infections, as well as empirically estimated effective contact rates over the period of interest *β*_*t*_, *t* ∈ [*t*_1_, *t*_2_]. We then use estimated time-varying ATTs for the effective contact rate, 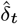 to impute the AME as follows:

1. Calculate the fitted treated potential outcomes as an average from 1000 infection trajectories simulated from an SEIR model with effective contact rates set to 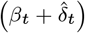, corresponding to an effective reproduction number 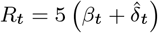.
2. Calculate the fitted untreated potential outcomes for the treated group as an average from 1000 infection trajectories simulated from an SEIR model with effective contact rate set to *β*_0_, corresponding to an effective reproduction number *R*_*t*_ = 5*β*_0_.
3. The AME for a log *R*_*t*_ or a log *β*_*t*_ model is then given by the average difference in projected infections over the post-intervention period between fitted trajectories: 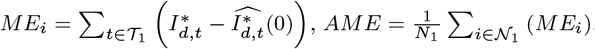.

## F. SEIR framework

We summarize the SEIR framework using the following equations:

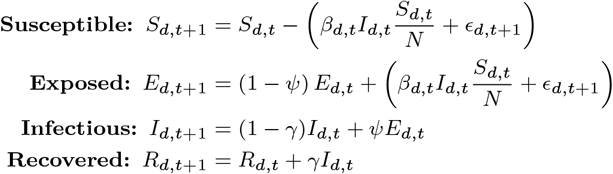

Compared with an SIR model specified in Eq. 1, the only additional parameter introduced here is *ψ*, which in our analysis is assumed to be a constant rate of infectious given exposure.

## G. Inference

We conduct inference using the wild score bootstrap, which allows for valid inference with heteroskedastic data and a small number of clusters when a generalized linear model is used for estimation. This is a generalization of the wild cluster bootstrap, proposed by Cameron et al. (23). The more widely-used wild cluster bootstrap perturbs the residual distribution for each bootstrap replicate based on a cluster-level random variable with mean 0 and variance 1, usually drawn from a Rademacher distribution (23, 24). Although this technique performs well with a small number of clusters, it requires a symmetric distribution in the residuals with mean 0, which is not satisfied by a Poisson generalized linear model (GLM). More broadly, this idea of perturbing distributions in bootstrap samples can be applied to the score contributions in the context of GLMs (23). Given any maximum likelihood estimation process, the score contribution for cluster *c* can be computed as the sum of score vectors in all observations from cluster *c*, where a score vector is the first derivative of the log-likelihood function. In each bootstrap replicate, we re-weight the score distribution based on an auxiliary cluster-level random variable with mean 0 and variance 1, and calculate a Wald statistic is calculated using the weighted scores. The p-value is the proportion of bootstrap replicates for which the bootstrapped Wald statistics exceed the observed Wald statistic under the null.

## Footnotes

* Astute readers may note that it is possible for 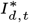 to exceed *S*_*d,t*_. This rarely occurs as typically *μ*_*t*_ << *S*_*d,t*_, but if a concern, the Poisson distribution can be substituted with a binomial.

† This framing is consistent with the practice of collapsing multiple time periods into a single step to address zero-valued outcomes when using log growth models.

‡ ”No intergenerational compounding” implies that individuals who become infectious at time *t* recover by the next time step *t* + 1.

§ Because *R* is traditionally used both to denote the “Recovered” individuals and the effective reproduction number, we follow this convention. In text that follows, *R* refers always to the effective reproduction number.

¶ There are some other approaches established in the literature to adjust when analyzing empirical data, such as inferring the unobserved initial generations of infections using a linear exponential growth model (30).

‖ Note that in the special case of constant exponential growth, 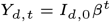, this condition reduces to 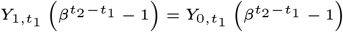.

